# The Two-dose MVA-BN Mpox Vaccine Induces a Nondurable and Low Avidity MPXV-specific Antibody Response

**DOI:** 10.1101/2024.01.28.24301893

**Authors:** Aaron L. Oom, Kesi K. Wilson, Miilani Yonatan, Stephanie Rettig, Heekoung Allison Youn, Michael Tuen, Yusra Shah, Ashley L. DuMont, Hayley M. Belli, Jane R. Zucker, Jennifer B. Rosen, Ramin Sedaghat Herati, Marie I. Samanovic, Ralf Duerr, Angelica C. Kottkamp, Mark J. Mulligan, the NYC OSMI Study Group

## Abstract

The 2022 global outbreak of clade IIb mpox was the first major outbreak of mpox outside of African nations. To control the outbreak, public health officials began vaccination campaigns using the third-generation orthopoxvirus vaccine MVA-BN. Prior to this outbreak, the durability of MPXV-specific immunity induced by MVA-BN was poorly understood. In 2022, we launched the New York City Observational Study of Mpox Immunity (NYC OSMI, NCT05654883), a longitudinal study of 171 participants comprising MVA-BN vaccinees and mpox convalescent individuals. Peripheral blood sampling was performed at intervals including prior to vaccination, after one dose, and after the second dose. MVA-BN vaccinees with and without a history of smallpox vaccination demonstrated detectable mpox virus (MPXV)-specific memory B cells at one-year post-vaccination. Additionally, MVA-BN increased MPXV neutralizing titers in smallpox vaccine-naïve vaccinees, with a comparable peak titer reached in naïve and smallpox vaccine-experienced vaccinees. However, neutralizing titers returned to baseline within 5-7 months for naïve individuals, while remaining elevated in those with prior smallpox vaccination. Both naïve and experienced individuals generated robust, immunodominant IgG responses against MPXV H3 and A35, but naïve vaccinees’ IgG responses showed lower avidity than experienced vaccinees. These data highlight a low avidity antibody response elicited by MVA-BN that is short-lived in naïve vaccinees. This work supports the need for long-term studies on protection induced by MVA-BN including the potential need for booster doses as well as the development of next-generation orthopoxvirus vaccines.

## Introduction

The 2022 global outbreak of clade IIb mpox (formerly known as monkeypox) was the first major outbreak of mpox outside of African nations with nearly 100,000 cases reported to date primarily in Europe and the Americas^1^. Over 34,000 cases have occurred in the US alone^2^. With cases primarily clustered among men who have sex with men, it was suggested that sexual transmission was playing a larger role in this outbreak^3^ compared to transmission from zoonosis or direct contact typically noted in endemic cases^4^. To stem rising case numbers worldwide, public health agencies launched awareness campaigns and began offering the two-dose MVA-BN vaccine series (Bavarian Nordic). MVA-BN (also known as IMVAMUNE, IMVANEX, or JYNNEOS) is an FDA-approved modified vaccinia Ankara vaccine indicated for prevention of mpox when administered twice subcutaneously (SC) over 28 days. MVA-BN is a third-generation non-replicating orthopoxvirus vaccine, in contrast to replication-competent vaccinia virus (VACV)-based first- and second-generation vaccines (e.g., DryVax or ACAM2000, respectively).

At the time of the 2022 outbreak, several studies had clearly established the decades of robust immunity generated by VACV vaccination^5–9^, but studies of MVA-BN durability had yielded more mixed results. Ilchman et al. found that VACV neutralizing titers wane to baseline levels within two years^10^, while Priyamvada et al. reported higher than baseline MPXV and VACV neutralizing titers at two years post-vaccination^11^. However, the latter study was conducted in the Democratic Republic of Congo where mpox is endemic, which may have impacted participant immunity. We still lack insight into key areas of antibody biology, including antibody specificities, avidity, and durability. Subsequent studies of the antibody repertoire following MVA-BN vaccination showed that antigen-specific antibodies were maintained at 2-3 months, but whether the titers would persist long-term remains unknown^12,13^.

To begin addressing knowledge gaps within MPXV-specific immunity following MVA-BN vaccination, we designed and conducted the New York City Observational Study of Mpox Immunity (NYC OSMI or OSMI). OSMI is a longitudinal study of MPXV-specific immunity in MVA-BN vaccinees and individuals with prior MPXV infection in NYC that began specimen collection in July 2022. We previously reported an interim analysis of OSMI where we observed a marked difference in binding antibody responses between MVA-BN vaccinees that had no prior smallpox vaccination (naïve vaccinees) and those that did (experienced vaccinees)^14^. Since then, there have been additional reports from other groups that further highlighted the differences between these two groups as well as the limited durability of MPXV- or VACV-specific binding and neutralizing antibodies in naïve MVA-BN vaccinees^15–18^.

In further characterization of these humoral responses, we observed that following vaccination the binding antibody titers that correlated with MPXV neutralization were low avidity and showed little to no avidity maturation by one-year post dose two in naïve vaccinees. However, both naïve and experienced vaccinees demonstrated detectable levels of MPXV-specific memory B cells, which may prove protective against subsequent exposures. These data highlight a nondurable, low avidity response from the two-dose regimen of MVA-BN in naïve people that warrants further study of booster doses^10,19^ and correlates of protection, more broadly.

## Methods

### Study Design

NYC OSMI investigates the MPXV-specific immune response in adults with and without HIV who have received MVA-BN vaccination as well as those who have recovered from mpox disease (NCT05654883, Institutional Review Board protocol 22-01338). Participants underwent sampling for serum, peripheral blood mononuclear cells (PBMCs), and saliva at specified intervals (Fig. 1A). Participants enrolled either pre- or post-MVA-BN vaccination, with the enrollment window extending up to 365 days post-first dose or mpox symptom onset. One hundred and seventy-one adult participants consented to be evaluated. Participants were recruited from city vaccination centers as well as by word of mouth. Cohort demographics and clinical information for participants analyzed here can be found in Table 1; additional details on mpox convalescent subjects are in Supplementary Table S1.

**Figure 1:**
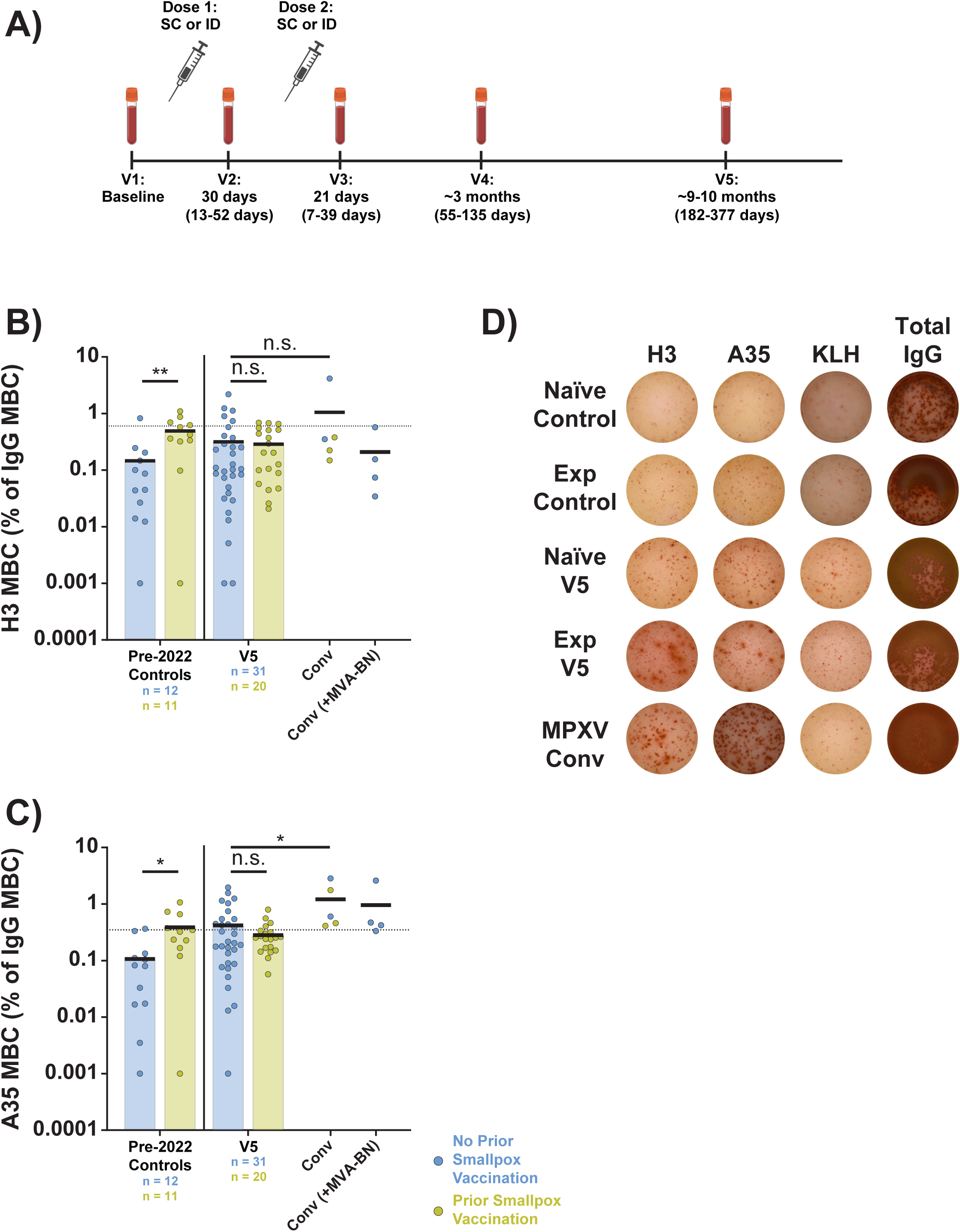
MPXV H3- and A35-specific memory B cells are detectable at one year after MVA-BN vaccination. IgG+ memory B cells (MBCs) were quantified via ELISpot. A) Diagram of study visits. For each visit the median days post-vaccination are shown along with the range of days used for each visit in parentheses. The windows of time denoted for each visit are used throughout the paper in analyses of specific visits. Diagram generated in BioRender. B) Aggregate MPXV H3-specific MBCs as a proportion of total IgG+ MBCs. C) Aggregate MPXV A35-specific MBCs as a proportion of total IgG+ MBCs. D) Representative well images from ELISpot assay. Convalescent (conv) samples were taken at ∼1 year post-mpox symptom onset. In panels B and C, colored/black bars indicate the mean. Black, dashed line in all panels is positivity threshold for binding as determined by 12 pre-2022 negative controls (mean of controls plus two times the standard deviation). Statistical testing conducted by Kruskal-Wallis test with Dunn’s method for multiple comparisons for panels B and C. Samples with a value of 0 have been displayed as 0.001% to accommodate the log scale; statistics were calculated with actual value. SC, subcutaneous; ID, intradermal; Exp, experienced vaccinees (prior history of smallpox vaccination); KLH, keyhole limpet hemocyanin (negative control).

**Table 1:** Demographics of the NYC Observational Study of Mpox Immunity.

|  | Total Participants |  | Prior Smallpox Vaccination (Experienced) |  | No Prior Smallpox Vaccination (Naïve) |  |
| --- | --- | --- | --- | --- | --- | --- |
|  | No. | % | No. | % | No. | % |
| **Enrolled Participants** | 159 |  | 49 | 30.8 | 110 | 69.2 |
| **Gender** |  |  |  |  |  |  |
| Male | 128 | 80.5 | 46 | 93.9 | 82 | 74.5 |
| Female | 18 | 11.3 | 3 | 6.1 | 15 | 13.6 |
| Non-binary/Gender Fluid/Queer | 13 | 8.2 | 0 | 0.0 | 13 | 11.8 |
| **Age** |  |  |  |  |  |  |
| Median | 38 |  | 62 |  | 33 |  |
| Range |  |  | 46 to 75 |  | 20 to 50 |  |
| **Race/Ethnicity** |  |  |  |  |  |  |
| White | 104 | 65.4 | 39 | 79.6 | 65 | 59.1 |
| Black or African American | 15 | 9.4 | 6 | 12.2 | 9 | 8.2 |
| Asian | 17 | 10.7 | 2 | 4.1 | 15 | 13.6 |
| Other | 23 | 14.5 | 2 | 4.1 | 21 | 19.1 |
| Non-Hispanic | 118 | 74.2 | 40 | 81.6 | 78 | 70.9 |
| Hispanic | 39 | 24.5 | 8 | 16.3 | 31 | 28.2 |
| Other | 2 | 1.3 | 1 | 2.0 | 1 | 0.9 |
| **LGBTQ+** |  |  |  |  |  |  |
| Yes | 142 | 89.3 | 46 | 93.9 | 96 | 87.3 |
| No | 17 | 10.7 | 3 | 6.1 | 14 | 12.7 |
| **Administration Route** |  |  |  |  |  |  |
| SC-SC | 35 | 22.0 | 10 | 20.4 | 25 | 22.7 |
| ID-ID | 31 | 19.5 | 13 | 26.5 | 18 | 16.4 |
| ID-SC | 16 | 10.1 | 8 | 16.3 | 8 | 7.3 |
| SC-ID | 66 | 41.5 | 16 | 32.7 | 50 | 45.5 |
| Incomplete/Unknown | 11 | 6.9 | 2 | 4.1 | 9 | 8.2 |
| **HIV Status** |  |  |  |  |  |  |
| PWH | 42 | 26.4 | 26 | 53.1 | 16 | 14.5 |
| HIV-uninfected | 117 | 73.6 | 23 | 46.9 | 94 | 85.5 |

Prior smallpox vaccination status was assessed through an adjudication process. For this process, the following factors were considered: physical examination of the upper arms for presence of scar consistent with past vaccinia vaccination, prior Bacillus Calmette-Guerin (BCG) vaccination, prior military service or other occupational risk requiring smallpox vaccination (including research laboratory work), country of birth, year of migration to the U.S., and whether a participant’s year of birth was consistent with active smallpox vaccine distribution in the participant’s country of origin. A literature review was performed to determine when individual countries ended their routine smallpox vaccination campaigns; e.g., 1972 in the US. Reviews of smallpox vaccination status were conducted by two infectious diseases physicians. For people with HIV (PWH), CD4+ T cell counts were obtained from electronic medical records at the time of enrollment (Supplementary Table S2). The date and route of mpox vaccination were verified through electronic medical records, participant’s description of the vaccination procedure, and/or records available through the New York City Health Department’s Citywide Immunization Registry.

Additional pre-2022 control samples and pre-OSMI enrollment participant samples were obtained from the NYU Langone Vaccine Center Biorepository (Institutional Review Board protocol 18-02035).

### Blood Collection

Venous blood was collected by standard phlebotomy. PBMCs were isolated from CPT vacutainers (BD Biosciences) and processed within four hours of collection. PBMCs were viably cryopreserved and thawed later for assays. Sera were collected in SST tubes (BD Biosciences) and frozen immediately at −80°C.

### MPXV Virus Stock Propagation

All mpox virus work was conducted in a certified ABSL3 facility at NYU Grossman School of Medicine. MPXV clade IIb, lineage B.1, was obtained from BEI Resources (NR-58622); this virus was isolated in Massachusetts, USA during the 2022 outbreak. Working stocks of MPXV were generated by plaque purification in Vero E6 cells (ATCC #CRL-1586) followed by three passages of propagation in Vero E6 cells at a multiplicity of infection (MOI) of 0.01 for three days each at 37°C. Virus was harvested by freeze-thaw lysis of scraped cells followed by centrifugation at 1,000x*g* to remove cell debris. At each passage, stocks were purified over 36% sucrose in TNE buffer (Quality Biological #351-302-101) at 32,900x*g* for 80 min at 4°C. Purified MPXV stocks were sequence-verified.

### Immunofluorescence-based MPXV Microneutralization Assay

One day before MPXV infection, 1.5×10^4^ Vero E6 cells were plated in each well of a black 96-well assay plate in cell culture media (Dulbecco’s Modified Eagle Medium [DMEM] supplemented with penicillin/streptomycin, 2 mM L-glutamine, and 10% heat-inactivated fetal bovine serum [FBS]) and incubated at 37°C and 5% CO_2_. All participant sera were complement-inactivated prior to use by heating for 30 min at 56°C. Participant serum was 2-fold serially diluted in infection media (DMEM lacking sodium pyruvate supplemented with 2% heat-inactivated FBS) starting at a 2.5-fold dilution (final starting dilution is 5-fold following addition of an equal volume of virus) for 8-points; all dilutions were performed in triplicate. Prior to infection, serum dilutions were incubated with MPXV (sufficient for MOI ≈ 0.01) for 1 hr at 37°C. Cells were washed with 1x phosphate-buffered saline (PBS) then incubated with serum-virus mixtures for 42 hrs at 37°C and 5% CO_2_. Following infection, plates were submerged in 10% formalin (Fisher Scientific #SF984) for 1 hr at room temperature (RT). Fixed samples were rinsed with water then permeabilized and blocked with 3% bovine serum albumin (BSA) and 0.1% Triton X-100 in PBS for 30 min at RT. Samples were then incubated with a polyclonal rabbit anti-vaccinia virus Lister strain antibody (Abbexa #abx023200) diluted 1:1,000 in 3% BSA in PBS (blocking buffer) for 1 hr at RT. All plates were washed four times with PBS then stained with 1:2,000 dilution of donkey anti-rabbit AlexaFluor647 secondary (Thermo #A-31573) and 1.25 µg/mL DAPI in blocking buffer. Following staining, cells were washed four times with 1x PBS before filling each well with 100 µL of PBS for imaging and quantification using the BioTek Cytation 7 Cell Imaging Multi-Mode Reader and Gen5 Image Prime software. MPXV neutralizing titers (ID_50_ values) were calculated from percent inhibition using GraphPad Prism 10.0.03 non-linear regression (variable slope with four parameters) with top and bottom constraints (100 and 0, respectively). Samples with ID_50_ values beyond the initial dilution range were repeated with an adjusted dilution curve.

### Multiplexed Immunoassay for Binding Antibodies and Avidity

Binding and avidity of MPXV-specific IgG in serum were measured using the Luminex platform for a 12-plex assay. Eight different MPXV proteins were included: A29 (Sino Biological #40891-V08E), A30 (Cell Sciences #YVV15001A), A35 (Sino Biological #40886-V08H), B16 (Cell Sciences #YVV17401A), B21 (Cell Sciences #YVV16301A), E8 (Cell Sciences #YVV13201A), H3 (Sino Biological #40893-V08H1), and L1 (Sino Biological #40889-V07E). In addition, quality control beads were included: rubella virus (RUBV) E1/E2 (ACROBiosystems #GL2-R5583), BSA (Thermo Scientific #J65097.22), human recombinant IgG1 (Sigma-Aldrich #I5154), and IC45 (Luminex #MRP1-045-01). Differing bead regions were coupled to antigen using the xMAP Antibody Coupling Kit (Luminex #40-50016) at the following optimized concentrations: 10 pmol/1×10^6^ beads for H3, A35, RUBV E1/E2, and human IgG1; 50 pmol/1×10^6^ beads for E8 and A29; and 100 pmol/1×10^6^ beads for A30, B16, B21, L1, and BSA. All MPXV antigens were His-tagged, and coupling was confirmed using an anti-His antibody (Abcam #ab27025). Beads were multiplexed with 1,000 beads per region for each sample.

Samples were analyzed by first incubating equal volumes of complement-inactivated serum (heated for 30 min at 56°C) and multiplexed beads for 1 hr in the dark on a shaker at RT. All sera were analyzed at final dilutions of 1:100 and 1:2,000, prepared in technical duplicate in 1x PBS (-Ca^2+^ -Mg^2+^) with 0.01% BSA and 0.02% Tween-20 (PBS-TB). The beads were then washed twice with PBS-TB before being incubated with either PBS (-Ca^2+^ -Mg^2+^) or the chaotropic agent, 2M ammonium thiocyanate (NH_4_SCN) for 30 min in the dark on a shaker at RT; one technical replicate of each dilution was used for each condition. Beads were washed again and then incubated for 30 min as before with rabbit anti-human IgG H&L detection antibody (Abcam #ab97158) diluted to 2 µg/mL in PBS-TB. Finally, the beads were washed before a final incubation as before with streptavidin-PE (BioLegend #405204) diluted to 4 µg/mL in PBS-TB for 30 min. The beads were washed a final time and then run on a Luminex 200 instrument with sample volume set to 50 µL. Instrument settings were as follows: 50 bead minimum per region and DD gate set 6,000 to 17,000. Binding titers were calculated by measuring the area under the curve (AUC) of the median fluorescence intensity of the two dilutions that received PBS, not NH_4_SCN. To calculate avidity, the binding AUC from the NH_4_SCN condition was divided by the AUC from the PBS condition to determine a ratio that is reported as the avidity index. Samples were normalized by the inclusion of a positive control pool on each plate, generated from pre-2022 controls with prior smallpox vaccination; normalization was conducted on an antigen-by-antigen basis.

### Anti-MPXV H3 Enzyme-Linked Immunosorbent Assay (ELISA)

Binding antibodies against MPXV H3 protein were measured via direct ELISA of participant serum as previously described^14^. In brief, 96-well plates were coated with 0.5 µg/mL MPXV H3 protein (Sino Biological Inc., 40893-V08H1) diluted in PBS. Plates were blocked with PBS +0.05% Tween 20 (Thermo Fisher Scientific) (PBS-T) +5% non-fat milk (blocking buffer) for 1 hr at RT. Heat-inactivated serum samples were serially diluted in blocking buffer and incubated in wells for 2 hrs at RT. Horseradish peroxidase-conjugated goat-anti-human IgG (Southern BioTech, 2040-05) was diluted in blocking buffer (1:2,000) and added to each well to incubate for 1 hr at RT. Finally, plates were developed for 5 min with 3,3’,5,5’-Tetramethylbenzidine Peroxidase Substrate (Thermo Fisher Scientific) followed by 1N hydrochloric acid to halt the assay. Absorbance at 450 nm was measured on a Synergy 4 (BioTek) plate reader. A reaction positivity cutoff was determined by calculating 2 times the standard deviation plus the mean of 16 pre-2022 control samples with no prior history of smallpox vaccination. Endpoint titers were calculated by interpolating at which reciprocal dilution a sample’s curve crossed the cutoff using the non-linear fit analysis in GraphPad Prism 9.5.1. Normalization for batch effect was addressed by the inclusion of a positive control pool generated from pre-2022 controls with prior smallpox vaccination. Sera with titers below the limit of detection (50) were scored as 25.

### Memory B Cell ELISpots

Memory B cells were quantified as previously described^20^ with application-specific changes elucidated here. Briefly, cryopreserved PBMCs were thawed and resuspended in 1 mL of warmed RPMI-1640 media supplemented with 10% heat-inactivated FBS and 2 mM L-glutamine (R10 media). Cells were counted and resuspended at 1×10^6^ cells/mL in stimulation media (R10 supplemented with 100 U/mL of DNase I, 1:1,000 dilution of B-Poly-S [Cellular Technology Limited #CTL-HBPOLYS-200], and 1 µM β-mercaptoethanol). A total of 2×10^6^ cells were added across 2 wells of a 24-well plate and incubated for 5 days at 37°C and 5% CO_2_. At least 18 hrs prior to day 6, Millipore 96-well multiscreen HA filter plates (Millipore #MSHAN4B50) were coated with 2 µg/mL of recombinant MPXV H3 or A35, 10 µg/mL of donkey anti-human IgG Fcγ fragment (Jackson ImmunoResearch # 709-005-098), or 4.16 µg/mL of Imject Mariculture keyhole limpet hemocyanin (KLH) (Thermo Scientific #77600), then incubated at 4°C. On day 6, coated assay plates were washed four times with 1x PBS-T then blocked with R10 media at 37°C and 5% CO_2_ for 1-2 hrs. During the blocking, stimulated PBMC cultures were transferred to 15 mL conical tubes, washed with warmed R10 media, then resuspended in R10 media at a concentration of 1×10^7^ cells/mL. A dilution series was prepared for each sample by adding 5×10^5^ cells to the first row of wells on the blocked assay plates, then serially diluting 3-fold for a 4-point dilution curve. For the total IgG plate, only 5×10^4^ cells were added. Plates were incubated for 6 hrs at 37°C and 5% CO_2_. Following incubation, plates were washed twice with PBS then four times with PBS-T. Plates were then incubated overnight at 4°C with biotinylated donkey anti-human IgG Fcγ fragment (Jackson ImmunoResearch # 709-065-098) diluted in PBS-T +2% FBS (antibody diluent). The next morning plates were washed four times with PBS-T then incubated with avidin-D-HRP (Vector Laboratories # A-2004-5) for 1 hr at RT. Plates were washed three times with PBS-T then three times with PBS before developing for five minutes with AEC substrate (BD Biosciences #551951). Following development, plates were washed twice with distilled water then allowed to dry for ∼24 hrs before imaging and spot counting on an ImmunoSpot automated ELISpot counter (Cellular Technology Limited). Samples with less than ∼750 IgG+ memory B cells per million PBMCs were removed from the analysis. Additionally, samples with KLH signal more than three times the standard deviation over the mean KLH signal were removed as outliers.

### Statistical Methods

Paired measurements in figures were analyzed by Wilcoxon matched-pairs signed rank test. Comparisons of multiple sample groups in figures were conducted by Kruskal-Wallis test with Dunn’s method for multiple comparisons. Linear mixed-effects regressions (LMER) were used to model neutralizing titer durability with repeated measurements. The *lme4* package (v.1.1-35.1) in R v.4.3.2 was used for all LMER calculations. Models were fit using the natural log transformation of the neutralizing titers. Both participant-level random intercepts only and participant-level random intercepts and slopes models were tested, but random intercepts only models were ultimately used as random intercepts and slopes models either failed to converge or had insufficient data for model fitting. Model predictions were generated using the *ggeffects* package (v.1.3.2). Half-lives were calculated by rearrangement of the LMER to its exponential decay form and use of the predicted slope for the days post dose 2 variable”. LMER model parameters for each instance are detailed in the Supplementary Materials. Correlation testing, paired measurements, and multiple comparison testing were all conducted in GraphPad Prism v.10.0.3. For all figures, p-value <0.05 is *, <0.01 is **, and <0.001 is ***.

## Results

### The New York City Observational Study of Mpox Immunity

We report here on 159 MVA-BN vaccinees with or without a prior history of smallpox vaccination (Table 1) and nine mpox convalescent individuals (Supplementary Table S1). The remaining three study participants were excluded due to missing data or visits. Study visit schedule is described in Figure 1A; participants could enroll at any time point. Possible time points included a baseline visit (V1), a visit between vaccine doses (V2), and three visits following receipt of the second vaccine dose: median visit dates of ∼3 weeks post dose two (V3), ∼3 months post dose two (V4), and ∼9-10 months post dose two (V5). Most participants were younger than 50 years old (70% of vaccinees) and identified as male and a member of the lesbian, gay, bisexual, transgender, and queer+ (LGBTQ+) community (77%), in line with the groups recommended to receive vaccination by the CDC^21^. Of note, the median age for vaccinees with a history of smallpox vaccination was 62, compared to 33 for those without prior smallpox vaccination (referred to henceforth as naïve participants). Nearly a third of the participants had previously received a smallpox vaccination (referred to henceforth as experienced participants).

Approximately half of those experienced participants were people with HIV (PWH), while only ∼15% of naïve participants were PWH. During the 2022 outbreak, limitations in available doses led to an emergency use authorization (EUA) for a dose-sparing measure involving intradermal (ID) delivery of one-fifth of the dose used SC^22,23^. Although our study includes a range of dosing combinations: SC-SC, SC-ID, ID-SC, and ID-ID, SC-ID was the most common in the study (∼33% of experienced participants and ∼46% of naïve participants).

### MPXV H3- and A35-specific memory B cells are detectable at one year following MVA-BN vaccination

As previous generations of smallpox vaccines generated robust cellular memory responses, we first examined whether MVA-BN induced antigen-specific memory B cells at a late time point, V5 (Fig. 1). Representative MPXV proteins from the intracellular mature virion and extracellular enveloped virion forms were chosen, H3 and A35, respectively. Antigen-specific IgG+ memory B cells were assayed by ELISpot in MVA-BN vaccinees as well as MPXV convalescent subjects. The proportions of vaccinees with positive levels (i.e., greater than background) of MBCs against H3 were 16% and 15% for naïve and experienced participants, respectively (Fig. 1B), and were 39% and 20% for A35, respectively (Fig. 1C). These data indicate durable establishment of MPXV-specific memory B cells in both cohorts in the study.

### Peak MPXV neutralization titers are comparable for naïve and experienced MVA-BN vaccinees but differ in durability

While MPXV-specific cellular responses may mitigate severe disease, antibodies may play an important role in preventing infection, specifically neutralizing antibodies. To measure the neutralizing titers in the sera of OSMI participants, a fluorescence-based microneutralization assay was developed using authentic clade IIb MPXV. Pre-outbreak samples from experienced individuals had higher neutralizing titers at baseline compared to naïve people (Fig. 2A, pre-2022 controls), in line with prior work showing long-lasting durability for orthopoxvirus immunity^5–9^. Aggregate data showed no difference between naïve and experienced participants at V3 (geometric mean titers [GMTs] of 95 versus 127, respectively), highlighting similar peak neutralizing titers (Fig. 2A). However, by V4, there was a difference between the two groups (GMTs of 45 for naïve versus 111 for experienced). This difference persisted at V5 with 81% of naïve vaccinees below the positivity threshold, compared to only 14% of experienced vaccinees (GMTs of 24 versus 85, respectively). Nine mpox-convalescent individuals were included as another control group, some of whom had received at least one dose of MVA-BN (Supplementary Table S1). At ∼1 year after symptom onset, these participants had GMTs comparable to peak titers (V3) for vaccinees, suggesting a more durable neutralizing antibody response post-MPXV infection as compared to post-MVA-BN vaccination. These trends in neutralizing antibodies were also observed for anti-MPXV H3 IgG binding antibody titers, as measured by ELISA, although a lower proportion of naïve MVA-BN vaccinees (50%) were below the limit of detection at V5 (Supplementary Fig. S1A). Taken together, these data suggest that while MVA-BN induces appreciable neutralizing titers in all vaccinees, the durability of these antibodies is limited in naïve vaccinees.

**Figure 2:**
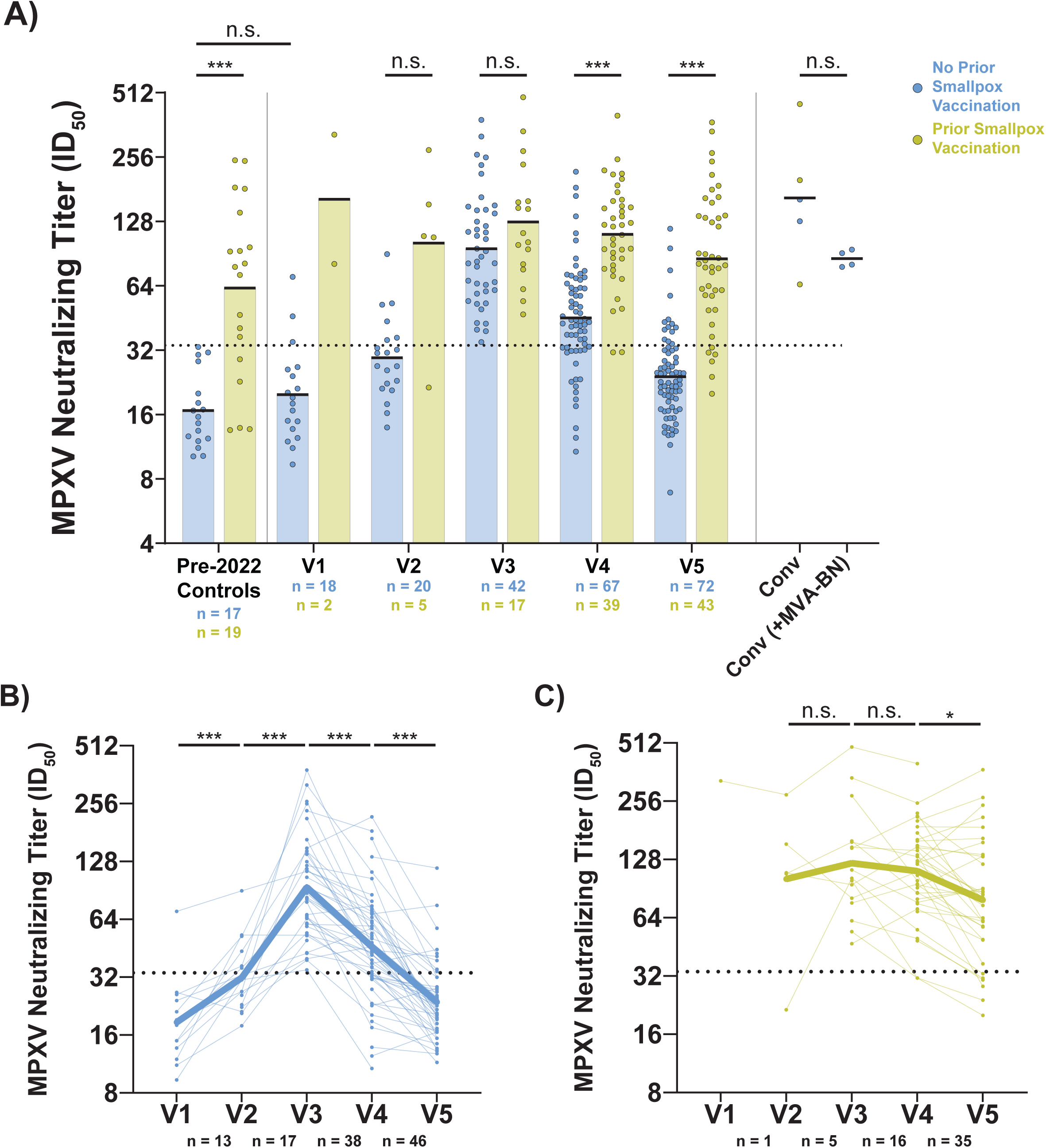
The MPXV neutralizing titers of smallpox vaccine-naïve individuals are less durable than those of experienced individuals following MVA-BN vaccination. A) Aggregate MPXV neutralizing titer data for each study visit sorted by prior smallpox vaccination status. Colored/black bars on each group indicate the geometric mean titer. Serum neutralizing titers were measured by a fluorescence-based microneutralization assay using authentic clade IIb MPXV. Pre-2022 controls come from the NYU Langone Vaccine Center Biorepository. Convalescent (conv) participants are sorted by whether they received MVA-BN following MPXV infection; all samples are taken at ∼1 year post-symptom onset. B) Longitudinal neutralizing titers from naïve participants across all five study visits. Bolded line indicates the mean neutralizing titer for participants at each time point. C) Longitudinal neutralizing titers from experienced participants across all five study visits. Bolded line indicates the mean neutralizing titer for participants at each time point. Horizontal, black dashed line in all panels indicates the positivity threshold for neutralization (ID_50_ = 33.7) as based on the pre-2022 negative controls (same method as Fig. 1). Statistical testing conducted by Kruskal-Wallis test with Dunn’s method for multiple comparisons for panel A and Wilcoxon matched-pairs signed rank test for panels B and C.

### HIV infection has no effect on responses to MVA-BN in people with CD4 counts over 250 cells/mm^3^

An important question in PWH is whether the EUA dose-sparing ID route was immunogenically comparable to the licensed full-dose SC route, as prior studies of MVA-BN vaccination in PWH had examined only the approved SC route^24,25^. All our participants with HIV had CD4+ T cell counts over 250 cells/cmm and most were over 500 cells/cmm (Supplementary Table S2). Combining all routes of vaccination, neutralizing titers were similar between HIV-uninfected participants and PWH in both the naïve and experienced groups (Supplementary Fig. S2). When focusing only on those that received at least one ID dose, a similar result was found (Supplementary Fig. S3). We additionally examined the effect of HIV status in a linear mixed-effects regression and similarly found no effect on neutralizing titers in naïve (Supplementary Table S3) or experienced participants (Supplementary Table S4). Lastly, using a linear mixed-effects model, we tested whether CD4 counts had an effect on neutralizing titers and found no such effect in naïve (Supplementary Table S5) or experienced participants (Supplementary Table S6). These data indicate that the MVA-BN vaccine established similar humoral responses in PWH in the settings of SC and reduced-dose ID vaccination.

### Neutralizing titers decay faster in naïve vaccinees compared to experienced vaccinees

To further leverage the longitudinal nature of the OSMI study, we next examined participants with samples from consecutive visits (Fig. 2B and C). Following the first dose of MVA-BN, naïve participants already had higher neutralizing titers relative to pre-vaccination (Fig. 2B).

Neutralizing titers peaked at V3 and declined thereafter. For experienced individuals (Fig. 2C), titers did not begin to decline until after V4. A similar pattern was seen with anti-MPXV H3 IgG titers (Supplementary Fig. S1B and C). To evaluate the rate of change in each group, paired fold changes in MPXV neutralizing titers for V3 versus V4 and V4 versus V5 were compared between naïve and experienced individuals. In both instances, naïve participants had a greater fold-change in titers from one visit to the next (Supplementary Fig. S4).

We then assessed this observation with statistical model, using linear mixed-effects regression (Supplementary Tables S3 and S4). For naïve participants, neutralizing titers were associated with dosing interval and days post dose two (model parameters can be found in Supplementary Table S7), while neutralizing titers in experienced participants were only associated with days post dose two (Supplementary Table S8). Using these models, the half-life of neutralizing antibodies was predicted to be 168 days (95% C.I.: 147 to 197 days) in naïve individuals, regardless of dosing interval. Participants that received the 1-month dosing interval, which is the minimum recommended interval, lost titer seropositivity by 160 days post dose two (Fig. 3A). The time to negativity increased by ∼1 month for each additional month between MVA-BN doses (Fig. 3A inset table). This was likely driven, in part, by the positive correlation of peak MPXV neutralizing titers with dosing interval (Supplementary Fig. 5). Antibody decline was far slower in experienced MVA-BN vaccinees (Fig. 3B) with a predicted half-life of 387 days (95% C.I.: 289 to 600 days). The predicted time to cross the seropositivity threshold was >350 days. Together these data highlight the difference in the longevity of serum neutralization activity for the two-dose MVA-BN regimen in naïve people versus experienced people.

**Figure 3:**
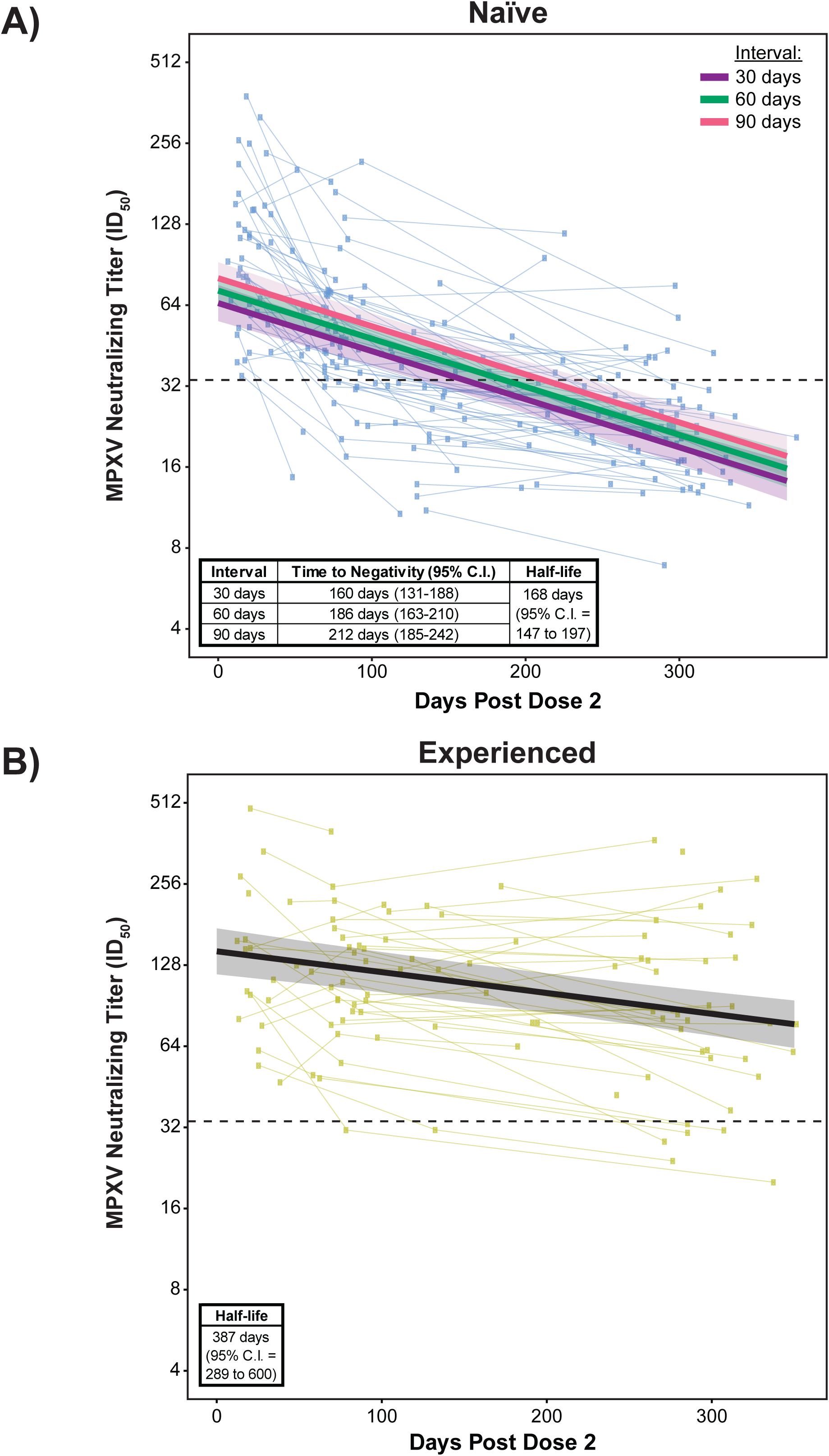
Modeling of neutralizing titer durability shows smallpox vaccine-naïve individuals revert to negative titers by ∼6 months, while experienced individuals remain positive beyond a year after MVA-BN vaccination. A) Linear mixed-effects regression of naïve participants using all data points after the second dose of MVA-BN. Regressions of natural log-transformed neutralizing titers use days post dose 2 and dosing interval as fixed effects and account for random subject effects by allowing for random intercepts. Colored regression lines with transparent ribbons indicate the predicted titers with 95% confidence interval. Model parameters are listed in Supplementary Table S7; 207 observations from 100 participants. B) Linear mixed-effects regression of experienced participants using all data points after the second dose of MVA-BN. Regression of natural log-transformed neutralizing titers uses days post dose 2 as a fixed effect and accounts for random subject effects by allowing for random intercepts. The solid black regression line with transparent ribbon indicates the predicted titers with 95% confidence interval. Model parameters are listed in Supplementary Table S8; 106 observations from 48 participants. Horizontal, black dashed line in each panel indicates the positivity threshold for neutralization (Fig. 2).

### IgG responses to MPXV A35 and H3 are immunodominant regardless of smallpox vaccination history, but have lower avidity in naïve vaccinees

To further characterize the serological responses of the naïve and experienced groups, a subset of participants from each group was chosen based on those that consistently participated from V3 to V5. These subsets had neutralizing titers similar to the full cohort (Supplementary Fig. S6). Using a multiplexed bead-based immunoassay, IgG binding titers and avidity for eight MPXV antigens (Table 2) were measured: A29, A30, A35, B16, B21, E8, H3, and L1. These surface antigens were selected as known protective targets^26–33^ or potential neutralizing targets based on previous reports^34–36^. IgG avidity was measured in this assay using an additional incubation with 2M ammonium thiocyanate following serum incubation. We found that the H3 and A35 viral proteins were immunodominant in both naïve and experienced individuals across all time points post-dose two (Supplementary Fig. S7A). Binding titers for H3 and A35 were also correlated with MPXV neutralization in both groups across multiple time points (Supplementary Fig. S7B). As the VACV homologs of H3 and A35 have been previously shown to be protective antibody targets in orthopoxvirus infections^26–28^, we decided to focus on these two antigens for our analysis.

**Table 2:** Eight MPXV proteins used in the multiplexed immunoassay for antibody binding and avidity: their location in virions and function in viral infection.

| **MPXV Protein** | **VACV Copenhagen Homolog** | **Function/Location** | **References** |
| --- | --- | --- | --- |
| H3 | H3 | Intracellular mature virus (IMV) protein, binds heparan | Ref. <sup>26,27</sup> |
| A35 | A33 | Extracellular enveloped virus (EEV) protein, envelope glycoprotein | Ref. <sup>28,32,33</sup> |
| E8 | D8 | IMV protein, binds chondroitin | Ref. <sup>29</sup> |
| B16 | B19 | Type I interferon antagonist | Ref. <sup>49</sup> |
| A29 | A27 | IMV protein, binds heparan | Ref. <sup>30,32</sup> |
| A30 | A28 | Member of entry-fusion complex | Ref. <sup>31</sup> |
| B21 | N/A | Presumed surface protein | Ref. <sup>36</sup> |
| L1 | J1 | IMV protein, virion morphogenesis | Ref. <sup>35,50</sup> |

Similar to MPXV neutralizing titers, both anti-H3 (Fig. 4A) and anti-A35 (Fig. 4B) IgG titers reached comparable peaks in naïve and experienced individuals. IgG titers against each antigen also declined faster in naïve participants as compared to experienced vaccinees. Anti-H3 IgG began to wane after V3 in both groups (Fig. 4C and D), while anti-A35 IgG began to wane after V3 in naïve individuals (Fig. 4E), but only after V4 in experienced individuals (Fig. 4F). The timing of A35 waning more closely matched the waning of neutralizing titers in experienced vaccinees. We also measured avidity for anti-H3 and anti-A35 IgG. Aggregate data showed higher IgG avidity for the H3 (Fig. 5A) and A35 proteins (Fig. 5B) in the experienced group across all time points post dose two. Surprisingly, naïve vaccinees had longitudinal changes in anti-H3 IgG avidity that, while statistically significant, were low and inconsistent across time (Fig. 5C). However, anti-H3 IgG avidity declined from V3 to V4 and V4 to V5 in the experienced group (Fig. 5D), perhaps indicating the generation of *de novo* low avidity responses that reduce the bulk avidity in the sera. Changes in the avidity of anti-A35 IgG were more subdued but in a similar direction (Fig. 5E and F). Taken together these data indicate similar B cell specificities (immunodominance) for naïve and experienced MVA-BN vaccinees, but with avidity being much higher in experienced MVA-BN vaccinees which may imply limited affinity maturation in naïve individuals.

**Figure 4:**
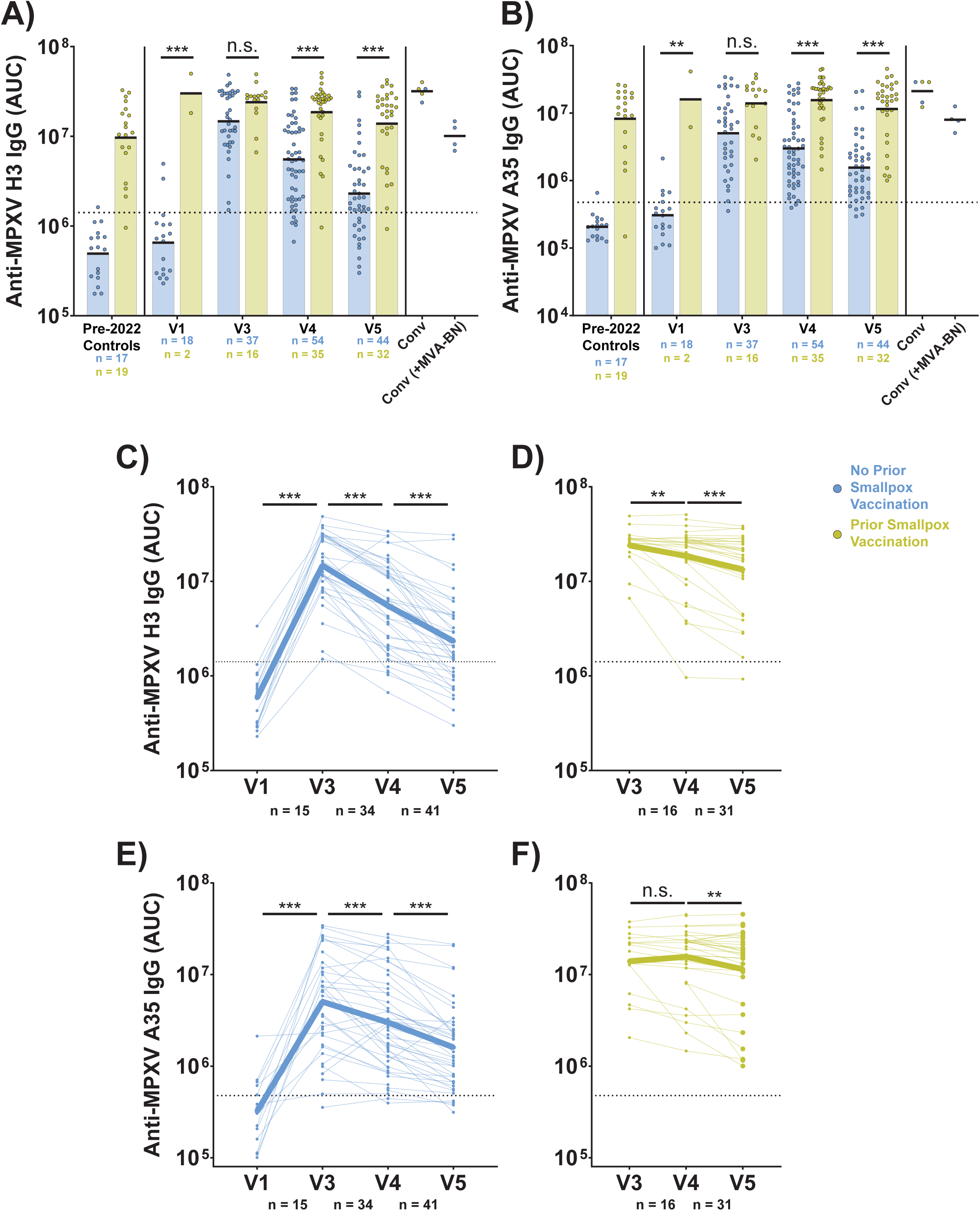
Anti-MPXV H3 and -A35 IgG reach comparable peak titers in smallpox vaccine-naïve and experienced participants, but wane more quickly in naïve participants following MVA-BN vaccination. A) Aggregate anti-MPXV H3 IgG titers as measured by multiplexed immunoassay. Titers are reported as AUC from two serum dilutions. B) Aggregate anti-MPXV A35 IgG titers as measured by multiplexed immunoassay. Titers are reported as AUC from two serum dilutions. C) Longitudinal anti-MPXV H3 IgG titers for naïve participants at V1 and V3-5 as measured by multiplexed immunoassay. Titers are expressed as AUC from two serum dilutions. D) Same as C, but for experienced participants across V3-5. E) Longitudinal anti-MPXV A35 IgG titers for naïve participants at V1 and V3-5 as measured by multiplexed immunoassay. Titers are expressed as AUC from two serum dilutions. F) Same as E, but for experienced participants across V3-5. MPXV convalescent (conv) samples were all taken at ∼1 year post-symptom onset. In panels A and B, colored/black bars indicate the geometric mean. For panels C-F, bolded line indicates the mean IgG titer for participants at each time point. Black, dashed line in all panels is positivity threshold for binding as determined by pre-2022 negative controls (same samples and method as in Fig. 2). Statistical testing conducted by Kruskal-Wallis test with Dunn’s method for multiple comparisons for panels A and B, and by Wilcoxon matched-pairs signed rank test for panels C-F.

**Figure 5:**
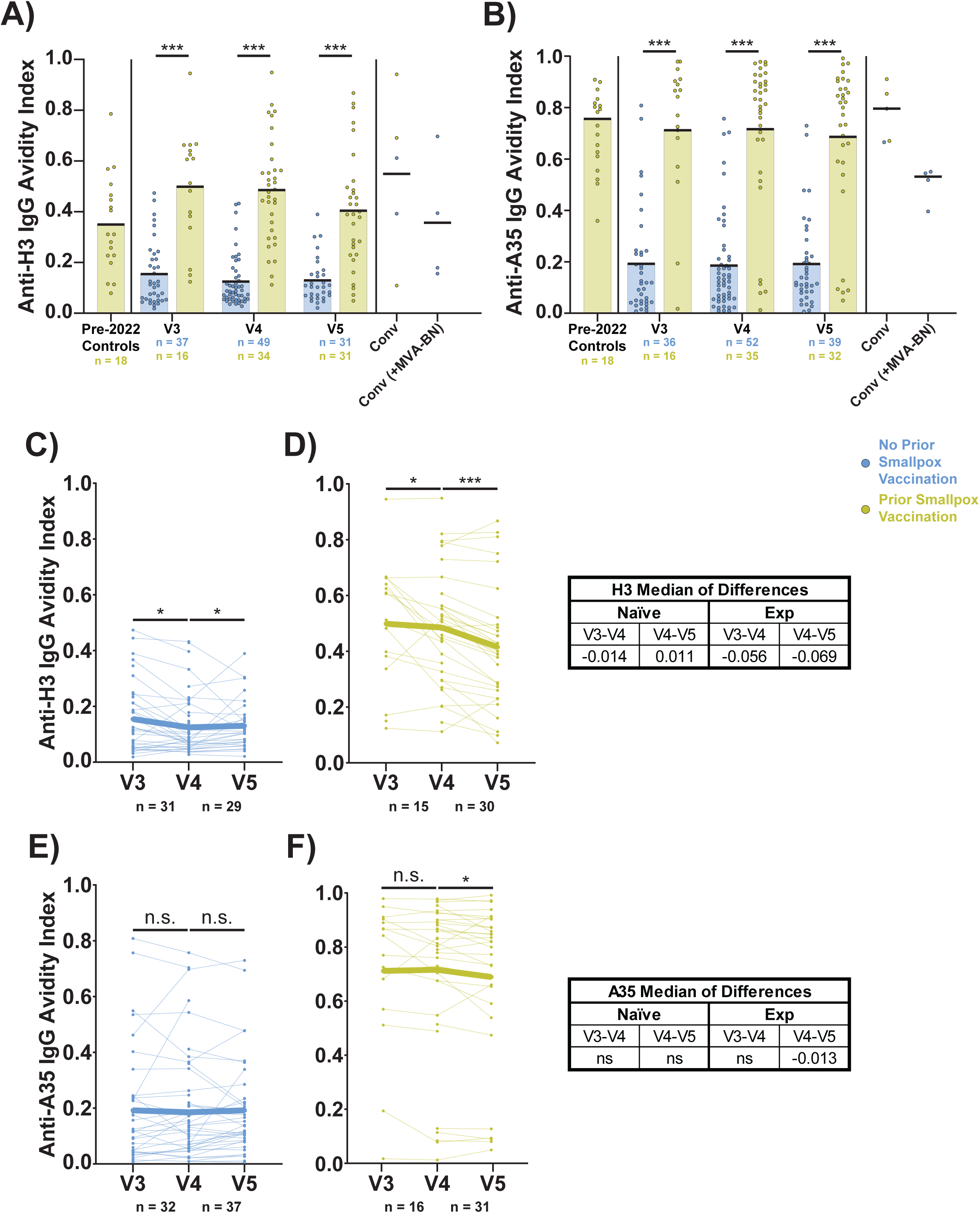
Smallpox vaccine-naïve vaccinees generate low avidity IgG against MPXV H3 and A35 after vaccination with MVA-BN. A) Aggregate avidity indices for anti-MPXV H3 IgG. B) Aggregate avidity indices for anti-MPXV A35 IgG. C) Longitudinal avidity indices for anti-MPXV H3 IgG antibodies for V3-5 of naïve participants. D) Same as C, but for experienced participants. E) Longitudinal avidity indices for anti-MPXV A35 IgG antibodies for V3-5 of naïve participants. F) Same as E, but for experienced participants. Avidity was measured in a multiplexed immunoassay using a 2M ammonium thiocyanate (NH_4_SCN) wash and expressed as the ratio of the AUC from the NH_4_SCN condition over the AUC from a PBS control condition. Convalescent (conv) samples were all taken at ∼1 year post-mpox symptom onset. In panels A and B, colored/black bars indicate the mean. Statistical testing conducted by Kruskal-Wallis test with Dunn’s method for multiple comparisons for panels A and B, and by Wilcoxon matched-pairs signed rank test for panels C-F. Exp, experienced vaccinees (prior history of smallpox vaccination).

## Discussion

We report here an analysis through one year of the MPXV-specific serum antibody magnitude, specificity, and avidity, and blood MBCs following the two-dose MVA-BN vaccination regimen. The current data indicate that IgG antibodies induced by MVA-BN are limited in their durability in naïve adults. We identified immunodominant MPXV proteins and found that the IgG response to those proteins was low avidity in naïve MVA-BN recipients in comparison to experienced MVA-BN recipients. We also demonstrated that MPXV-specific MBCs are detectable in circulation in a subset of vaccinees one year after vaccination despite the limited durability of circulating MPXV-specific antibodies.

The differences between naïve and experienced MVA-BN vaccinees may have several potential causes. While the experienced group had previously received a dose of replicating vaccine (vaccinia), the naïve group had received only the non-replicating MVA-BN. The two groups also differ in quantity of total vaccine doses (three versus two doses) as well as the timing of first vaccine exposure (childhood versus adulthood). To the former point, it is possible that an additional dose of MVA-BN would put the naïve group at parity with the experienced group, which should be investigated with booster studies. Related to the latter point, prior work has shown that first-time vaccination with 1^st^ or 2^nd^ generation replicating VACV vaccines in adulthood yields more durable neutralizing titers than were observed here in naïve adults receiving MVA-BN (∼10 years versus ∼6 months, respectively)^9^. This difference in initial poxvirus vaccination platform is likely to generate different inflammatory states, which we have recently shown durably impacts the CD4+ T cell memory response^37^. Taken together, these data would argue that VACV vaccine replication is a significant contributor to the durability of circulating antibody.

Beyond the impact of prior smallpox vaccination of MVA-BN responses, we observe an association between dosing interval and neutralizing titer durability. We previously reported no effect of dosing interval on anti-MPXV H3 IgG titers in naïve individuals^14^, which could imply differences in humoral responses for anti-H3 IgG binding titers and orthopoxvirus neutralization^38,39^. It is interesting to note that there was no effect of interval on neutralizing titers in experienced individuals. A possible explanation supported by previous work^40^ is that the second dose of MVA-BN provides no further immune boost in this group, but additional study is needed. The delayed rollout of second doses during the 2022 outbreak offered a unique opportunity to begin studying varied dosing schedules, but controlled studies will be necessary to identify an ideal dose interval and whether the relationship between dosing interval and titer durability remains linear beyond three months.

While we did find a proportion of vaccinees with H3- and A35-specific MBCs in circulation at one year after vaccination, these proportions were lower than the positivity rates of corresponding antibodies for each. As MBCs can also reside in tissues, such as the lymph node and spleen, this could diminish their availability for sampling via peripheral blood. Further, we saw no difference in MBCs between naïve and experienced vaccinees despite such a difference existing for antibody titers. As plasma cells (PCs) are the actual producers of antibodies, it is possible that there is a disconnect between PC formation and establishment of an MBC reservoir as these two populations are distinct. Further interrogation of the bone marrow compartment or germinal centers following MVA-BN vaccination is needed to better understand how PCs and MBCs are being formed. Such studies will also clarify what long-term protection is offered by MVA-BN vaccination.

These data are particularly timely in light of the ongoing mpox clade I outbreak in the Democratic Republic of Congo^41,42^. As vaccination remains limited in the DRC and neighboring nations, our data would support further clinical evaluation of several dose-sparing approaches such as a 1/5^th^ ID dose as well as delayed second doses. This clade I outbreak is also a critical opportunity to understand how MPXV immunity may differ between the two clades. As the two clades share ∼95% sequence identity^43^, it is possible that immunity is highly conserved.

As an observational study, OSMI is limited in a few ways. The emergent 2022 mpox outbreak limited collection of baseline samples for many of the participants in OSMI, especially the smaller group of individuals with prior smallpox vaccination. The speed of the public health emergency also created difficulties in defining narrow time windows for each study visit, as would be done in a typical vaccine clinical trial. In addition, childhood records of smallpox vaccination were incomplete. Beyond the limitations of observational studies on patient recruitment and classification, we report here only a limited view of cellular immunity. Future efforts should consider cellular immunity more broadly when studying differences established by replicating versus nonreplicating vaccine. In addition, study of antibody responses against MPXV proteins beyond those examined here may offer further insights into important neutralizing targets.

Directions for future work include further study of the magnitudes, specificities, qualities, and durability of memory B and T cells following the two-dose MVA-BN regimen. Our work demonstrates that MPXV-specific MBCs can remain in circulation at least a year following MVA-BN vaccination. However, a critical unanswered question is whether the cellular memory responses induced by the MVA-BN two-dose regimen in naïve vaccinees are sufficient to protect against mpox disease when circulating neutralizing antibody is no longer detectable. Further studies of breakthrough infections, booster doses, and correlates of protection are needed to better answer these questions. In addition, the immunity induced by MPXV infection will need further characterization to determine whether subsequent vaccination is needed for durable, protective immunity.

Overall, we have shown that the two-dose MVA-BN regimen in naïve individuals produces low avidity, nondurable serological responses, in contrast to the response in individuals with prior smallpox vaccination, whose antibody responses are durable and high avidity. An ideal future mpox vaccine would combine the safety advantages of the non-replicating MVA-BN vaccine with the robust, durable protective immunity of replicating VACV vaccines. Future studies of MVA-BN cellular immunity may offer clues as to how to improve immune responses, whether that is through adjuvant, additional doses, higher doses, altered dosing intervals, or even virologic changes to MVA-BN itself. This is particularly important as mpox outbreaks look poised to continue. We hope that these discoveries into antibody specificity, quality, and durability will offer crucial insights for the next-generation of mpox-specific vaccines^44–48^.

## Data Availability

All data produced in the present study are available upon reasonable request to the authors.

## Acknowledgements

Most importantly, the authors thank the NYC OSMI study participants for their altruism and volunteerism, without whom we would have no study. The authors also wish to acknowledge the full NYC OSMI Study Group, which includes, in addition to the byline authors, the following colleagues: Abdul Abdulai; Robert Arciuolo, MPH, CPH; Jaqueline Callahan, RN; Ellie Carmody, MD, MPH; Tamia Davis, NP; Amanda Dontino; Aimee Edwin, RN; Celia Engelson, NP; Andrew Fleming, MD; Olivia Frank, MPH; Emily Geesey; Shelby Goins; Victoria Guerra, RN; Erika Guevarra; Sarah Haiken; Aaliyah Henry; Kathryn Jano; Trishala Karmacharya; Dorothy Knutsen, MD; Irma Noriega, NP; Maria Null; Samuel Nweke; Lalitha Parameswaran, MD, MPH; Robert Pitts, MD, MPH; Gurchetan Randhawa, MD; Miguel Rodriguez; Pamela Suman; Apoorva Talanki; Meron Tasissa; Kathryn Swindell, NP; Julia Wagner, MPH; Jimmy P. Wilson; Samantha Yip, RN; Lisa Zhao. The authors would like to thank Dr. Meike Dittmann and Austin Schinlever for their assistance in cultivating MPXV viral stocks. We also thank the NYU Office of Scientific Research Animal Biosafety Level 3 staff, Drs. Ludovic Desvignes, Dominick Papandrea, and Alison Gilchrist, for their support of this work. Financial support was provided by the Blavatnik Family Foundation, the National Institutes of Health (NIH) grants no. AI148574 and 75N93021C00014 (to M.J.M.), the NYC Department of Health and Mental Hygiene, and NYU Grossman School of Medicine institutional support.

