## Supplemental Materials for "The Two-dose MVA-BN Mpox Vaccine Induces a Nondurable and Low Avidity MPXV-specific Antibody Response"

**Table S1: Sample information for convalescent participants.**

| Participant | Age Range | Prior Smallpox Vaccination | HIV Status | Received Tecovirimat? | Dose 1 DPO | Dose 1 Route | Dose 2 DPO | Dose 2 Route | Sample DPO |
|---|---|---|---|---|---|---|---|---|---|
| Conv 1 | 40-50 | No | PWH | Yes | -25 | SC | 46 | ID | 340 |
| Conv 2 | 30-40 | No | HIV uninfected | No | 9 | SC | 87 | ID | 336 |
| Conv 3 | 20-30 | No | HIV uninfected | Yes | -1 | SC | | | 386 |
| Conv 4 | 40-50 | No | PWH | Yes | 78 | ID | | | 388 |
| Conv 5 | 30-40 | No | PWH | Yes | | | | | 335 |
| Conv 6 | 50-60 | Yes | PWH | Yes | | | | | 338 |
| Conv 7 | 60-70 | Yes | PWH | No | | | | | 391 |
| Conv 8 | 60-70 | Yes | PWH | No | | | | | 433 |
| Conv 9 | 30-40 | No | PWH | No | | | | | 386 |

Participants above the horizontal line received one or two doses of MVA-BN.
Conv, convalescent; PWH, person with HIV; SC, subcutaneous; ID, intradermal; DPO, days post-symptom onset.

**Table S2: CD4 counts at time of enrollment for OSMI participants with HIV.**

| Naïve CD4 Counts | Exp CD4 Counts |
|---|---|
| 267 | 284 |
| 321 | 350 |
| 347 | 351 |
| 491 | 438 |
| 500.5 | 450 |
| 574.5 | 482.5 |
| 576 | 538.5 |
| 648 | 552 |
| 744 | 600.5 |
| 750 | 675 |
| 797 | 680 |
| 842 | 680 |
| 907 | 685 |
| 1387 | 703 |
| 1500 | 751.5 |
| | 755 |
| | 760 |
| | 859 |
| | 876 |
| | 890 |
| | 902 |
| | 948 |
| | 996 |
| | 1069 |
| | 1124 |

Exp, prior smallpox vaccination.

**Table S3: Linear mixed-effects model parameters for naïve participants using days post dose 2 (DPV2), age, HIV status, route of administration, and dosing interval as fixed effects and subject (OSMI) as a random effect; age is no longer significant following the removal of HIV status and route of administration.**

| **log(Neut)** | | | |
|---|---|---|---|
| *Predictors* | *Estimates* | *CI* | *p* |
| (Intercept) | 4.4587 | 3.9333 – 4.9842 | **<0.0001** |
| DPV2 | -0.0041 | -0.0047 – -0.0035 | **<0.0001** |
| Age | -0.0146 | -0.0281 – -0.0011 | **0.0347** |
| HIV [Pos] | 0.0068 | -0.2609 – 0.2745 | 0.9599 |
| Route [ID-SC] | 0.3527 | -0.0468 – 0.7522 | 0.0833 |
| Route [SC-ID] | 0.1622 | -0.0967 – 0.4211 | 0.2181 |
| Route [SC-SC] | 0.1504 | -0.1429 – 0.4438 | 0.3131 |
| Interval | 0.0030 | 0.0008 – 0.0052 | **0.0084** |
| **Random Effects** | | | |
| $\sigma^2$ | 0.17 | | |
| $\tau_{00\ OSMI}$ | 0.13 | | |
| ICC | 0.43 | | |
| $N_{OSMI}$ | 100 | | |
| Observations | 207 | | |
| Marginal $R^2$ / Conditional $R^2$ | 0.467 / 0.694 | | |
| REML Criterion at Convergence | 354.5 | | |

**Table S4: Linear mixed-effects model parameters for experienced participants using days post dose 2 (DPV2), dosing interval, route of administration, age, and HIV status as fixed effects and subject (OSMI) as a random effect.**

| **ln(Neut)** | | | |
|---|---|---|---|
| *Predictors* | *Estimates* | *CI* | *p* |
| (Intercept) | 4.1590 | 2.3366 – 5.9814 | **<0.0001** |
| DPV2 | -0.0018 | -0.0024 – -0.0011 | **<0.0001** |
| Age | 0.0142 | -0.0081 – 0.0366 | 0.2091 |
| HIV [Pos] | 0.1068 | -0.2439 – 0.4575 | 0.5469 |
| Route [ID-ID] | -0.0481 | -1.3567 – 1.2605 | 0.9420 |
| Route [ID-SC] | -0.4648 | -1.8034 – 0.8739 | 0.4923 |
| Route [SC-ID] | -0.1868 | -1.4623 – 1.0886 | 0.7718 |
| Route [SC-SC] | -0.2444 | -1.5097 – 1.0209 | 0.7023 |
| Interval | 0.0011 | -0.0032 – 0.0054 | 0.6144 |
| **Random Effects** | | | |
| $\sigma^2$ | 0.11 | | |
| $\tau_{00\ OSMI}$ | 0.30 | | |
| ICC | 0.74 | | |
| $N_{OSMI}$ | 48 | | |
| Observations | 106 | | |
| Marginal $R^2$ / Conditional $R^2$ | 0.153 / 0.779 | | |
| REML Criterion at Convergence | 187.6 | | |

**Table S5: Linear mixed-effects model parameters for naïve PWH using days post dose 2 (DPV2) and CD4 counts as fixed effects and subject (OSMI) as a random effect.**

| **ln(Neut)** | | | |
|---|---|---|---|
| *Predictors* | *Estimates* | *CI* | *p* |
| (Intercept) | 4.4747 | 4.0205 – 4.9288 | **<0.0001** |
| DPV2 | -0.0036 | -0.0049 – -0.0023 | **<0.0001** |
| CD4 | -0.0003 | -0.0008 – 0.0002 | 0.2310 |
| **Random Effects** | | | |
| $\sigma^2$ | 0.10 | | |
| $\tau_{00\ OSMI}$ | 0.05 | | |
| ICC | 0.36 | | |
| $N_{OSMI}$ | 14 | | |
| Observations | 29 | | |
| Marginal $R^2$ / Conditional $R^2$ | 0.532 / 0.700 | | |
| REML Criterion at Convergence | 53.3 | | |

**Table S6: Linear mixed-effects model parameters for experienced PWH using days post dose 2 (DPV2) and CD4 counts as fixed effects and subject (OSMI) as a random effect.**

| ln(Neut) | | | |
|---|---|---|---|
| *Predictors* | *Estimates* | *CI* | *p* |
| (Intercept) | 4.5443 | 3.6179 – 5.4707 | **<0.0001** |
| DPV2 | -0.0016 | -0.0025 – -0.0008 | **0.0004** |
| CD4 | 0.0006 | -0.0006 – 0.0019 | 0.3100 |
| **Random Effects** | | | |
| $\sigma^2$ | 0.08 | | |
| $\tau_{00\ OSMI}$ | 0.44 | | |
| ICC | 0.84 | | |
| $N_{OSMI}$ | 25 | | |
| Observations | 52 | | |
| Marginal $R^2$ / Conditional $R^2$ | 0.095 / 0.856 | | |
| REML Criterion at Convergence | 105.1 | | |

**Table S7: Linear mixed-effects model parameters for naïve participants using days post dose 2 (DPV2) and dosing interval as fixed effects and subject (OSMI) as a random effect.**

| ln(Neut) | | | |
|---|---|---|---|
| *Predictors* | *Estimates* | *CI* | *p* |
| (Intercept) | 4.0699 | 3.8736 – 4.2662 | **<0.0001** |
| DPV2 | -0.0041 | -0.0047 – -0.0035 | **<0.0001** |
| Interval | 0.0036 | 0.0014 – 0.0057 | **0.0011** |
| **Random Effects** | | | |
| $\sigma^2$ | 0.17 | | |
| $\tau_{00\ OSMI}$ | 0.13 | | |
| ICC | 0.44 | | |
| $N_{OSMI}$ | 100 | | |
| Observations | 207 | | |
| Marginal $R^2$ / Conditional $R^2$ | 0.449 / 0.692 | | |
| REML Criterion at Convergence | 344.6 | | |

**Table S8: Linear mixed-effects model parameters for experienced participants using days post dose 2 (DPV2) as a fixed effect and subject (OSMI) as a random effect.**

| ln(Neut) | | | |
|---|---|---|---|
| *Predictors* | *Estimates* | *CI* | *p* |
| (Intercept) | 4.9694 | 4.7733 – 5.1656 | **<0.0001** |
| DPV2 | -0.0018 | -0.0024 – -0.0012 | **<0.0001** |
| **Random Effects** | | | |
| $\sigma^2$ | 0.11 | | |
| $\tau_{00\ OSMI}$ | 0.29 | | |
| ICC | 0.73 | | |
| $N_{OSMI}$ | 48 | | |
| Observations | 106 | | |
| Marginal $R^2$ / Conditional $R^2$ | 0.091 / 0.754 | | |
| REML Criterion at Convergence | 170.2 | | |

Figure S1

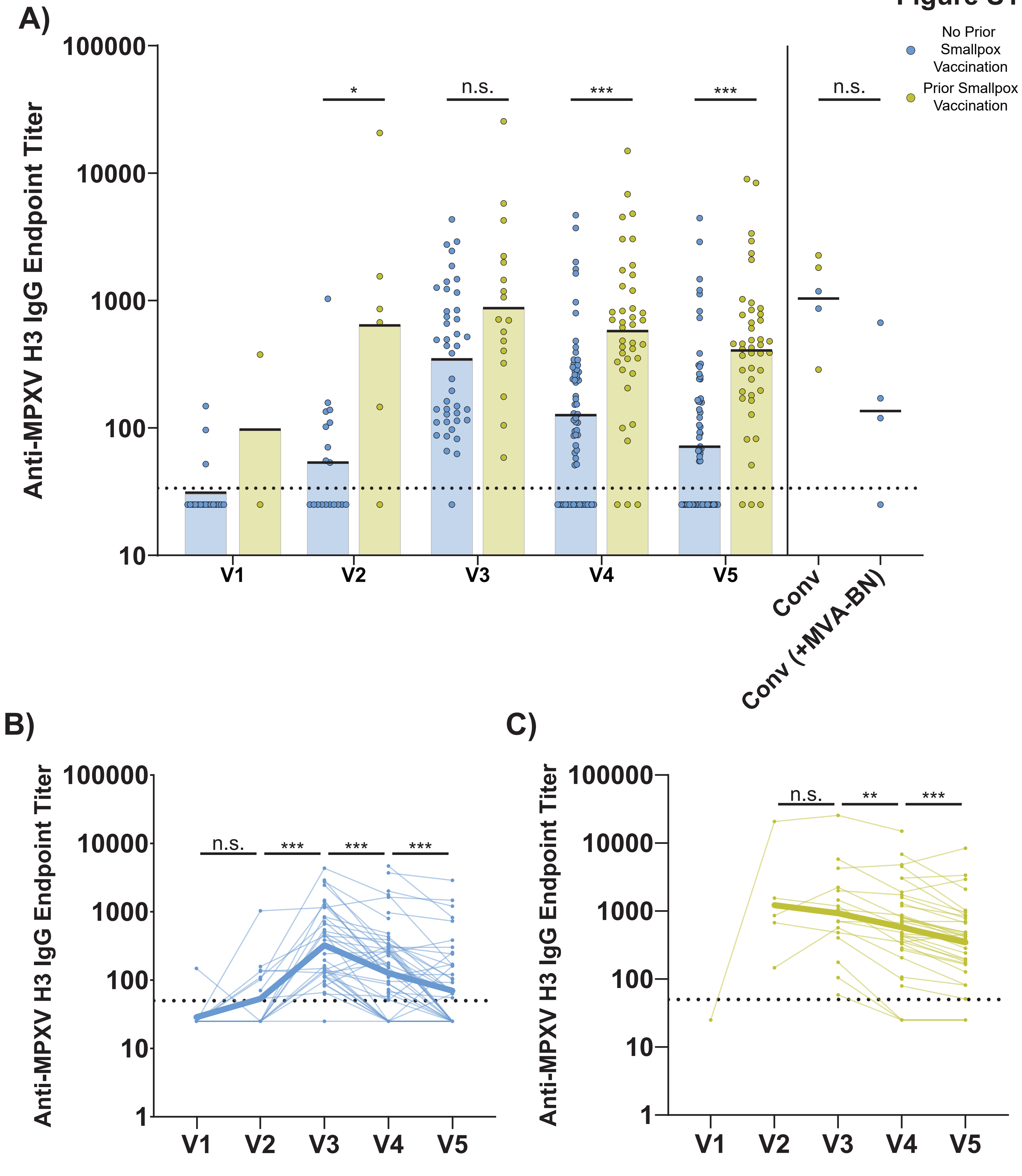
A)
Anti-MPXV H3 IgG Endpoint Titer
100000
10000
1000
100
10
*
n.s.
***
***
n.s.
V1
V2
V3
V4
V5
Conv
Conv (+MVA-BN)
No Prior Smallpox Vaccination
Prior Smallpox Vaccination
B)
Anti-MPXV H3 IgG Endpoint Titer
100000
10000
1000
100
10
1
n.s.
***
***
***
V1
V2
V3
V4
V5
C)
Anti-MPXV H3 IgG Endpoint Titer
100000
10000
1000
100
10
1
n.s.
**
***
V1
V2
V3
V4
V5

**Figure S1: Anti-MPXV H3 binding titers follow the general trends of MPXV neutralizing titers.** A) Serum anti-MPXV H3 IgG titers were measured by ELISA across all participants and study visits. Colored/black bars on each group indicate the geometric mean titer. Horizontal, dashed black line indicates the limit of detection (endpoint titer of 50) for the assay. Any values below the LoD are denoted as an endpoint titer of 25. B and C) Paired anti-MPXV H3 IgG endpoint titers by visit for naïve (B) or experienced (C) participants. Bolded line in panels B and C indicates the mean titer at each time point. Statistical testing conducted by Kruskal-Wallis test with Dunn’s method for multiple comparisons for panel A, and by Wilcoxon matched-pairs signed rank test for panels B and C.

### Figure S2

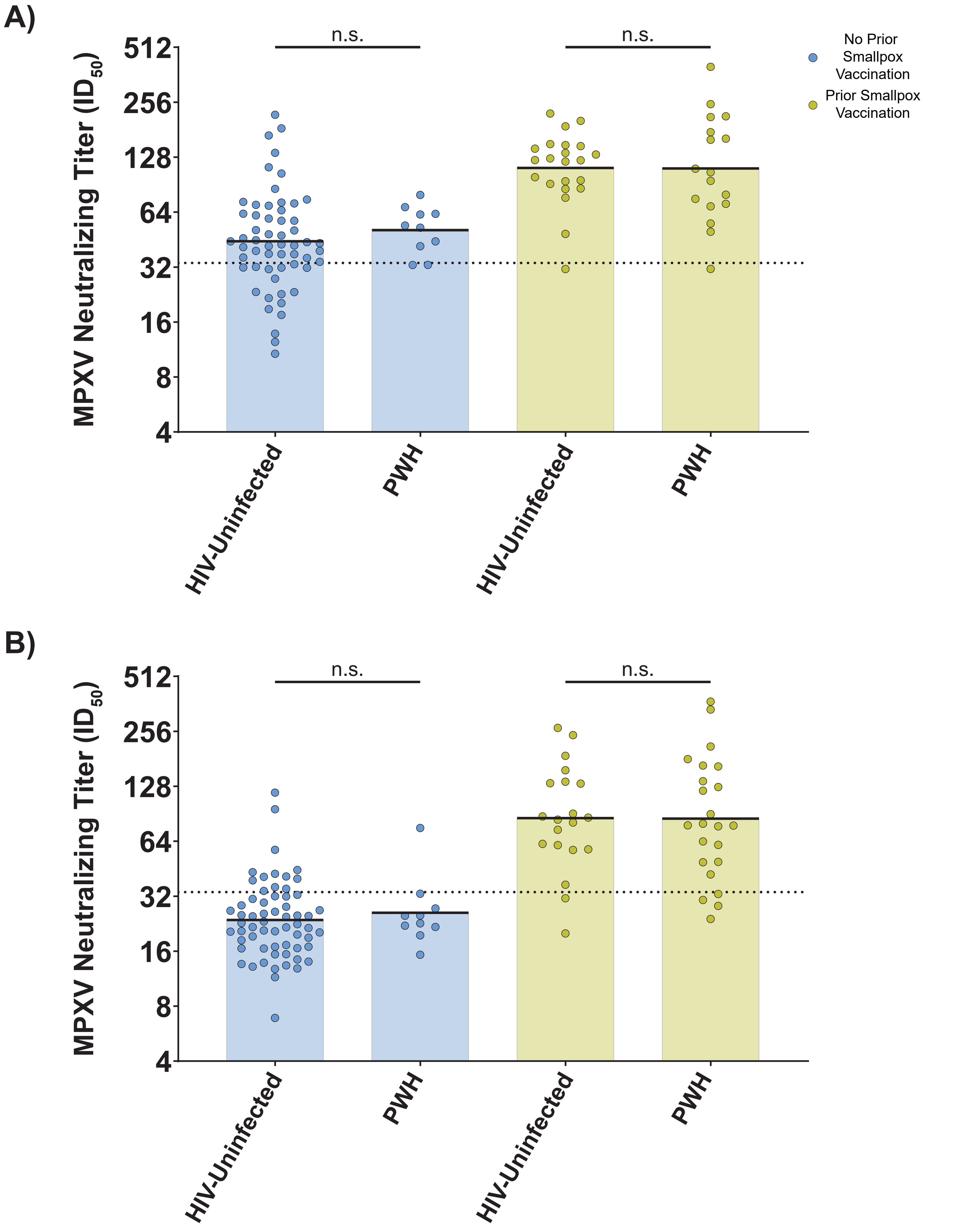
A)
n.s.
n.s.
512
256
128
64
32
16
8
4
MPXV Neutralizing Titer ($ID_{50}$)
HIV-Uninfected
PWH
HIV-Uninfected
PWH
No Prior Smallpox Vaccination
Prior Smallpox Vaccination
B)
n.s.
n.s.
512
256
128
64
32
16
8
4
MPXV Neutralizing Titer ($ID_{50}$)
HIV-Uninfected
PWH
HIV-Uninfected
PWH

**Figure S2: Neutralizing titers elicited by MVA-BN are statistically similar between PWH and HIV-uninfected individuals.** A) Aggregate MPXV neutralizing titer data for V4 stratified first by prior smallpox vaccination then by HIV status. Colored/black bars on each group indicate the geometric mean titer. Serum neutralizing titers were measured by a fluorescence-based microneutralization assay using authentic clade IIb MPXV. Horizontal, black dashed line in each panel indicates the positivity threshold for neutralization (Fig. 2). B) Same as A, but for V5. Statistical testing conducted by Kruskal-Wallis test with Dunn's method for multiple comparisons.

### Figure S3

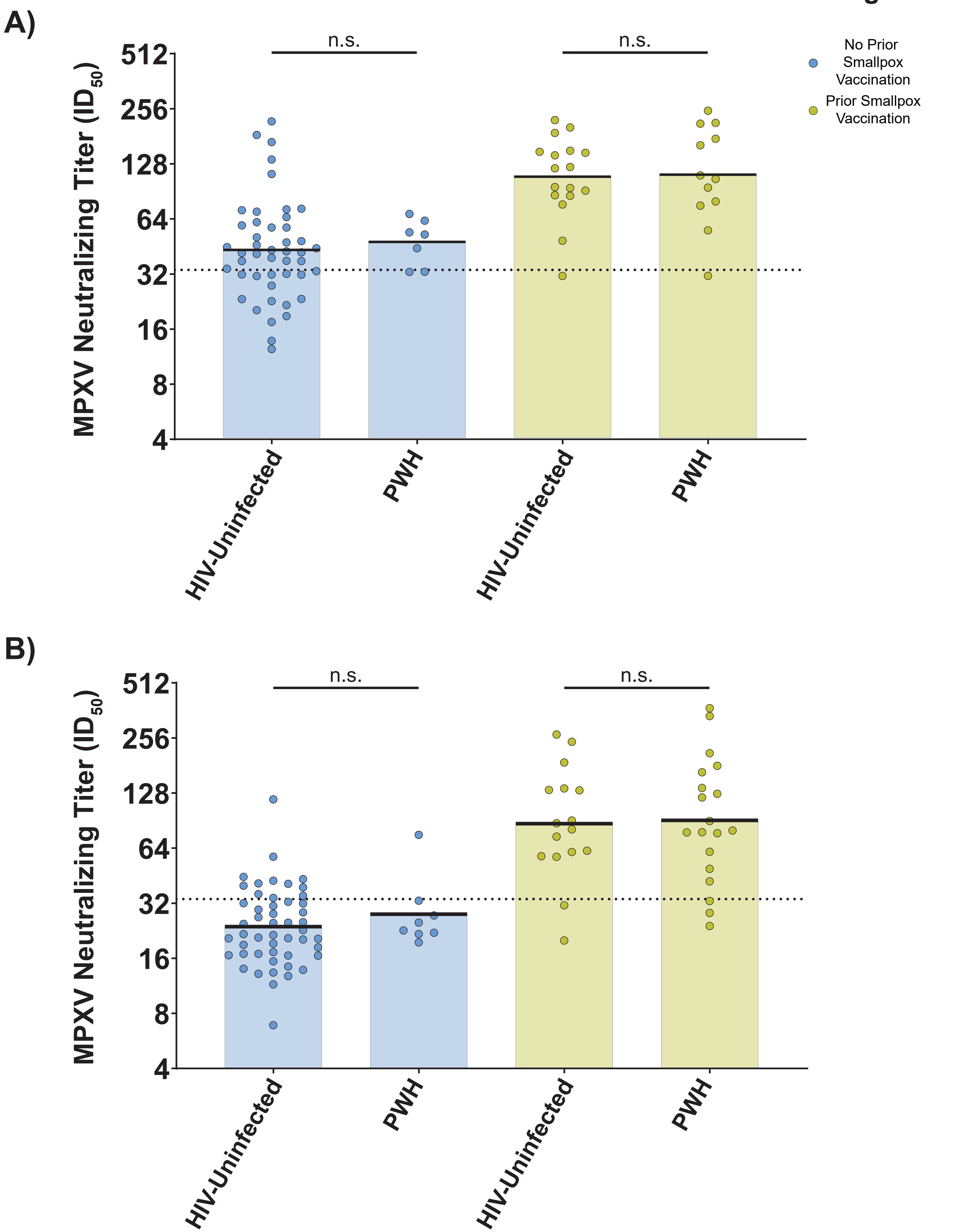

A)
B)
n.s.
n.s.
512
256
128
64
32
16
8
4
MPXV Neutralizing Titer (ID50)
HIV-Uninfected
PWH
HIV-Uninfected
PWH
No Prior Smallpox Vaccination
Prior Smallpox Vaccination

**Figure S3: Neutralizing titers elicited by intradermal MVA-BN are statistically similar between PWH and HIV-uninfected individuals.** A) Aggregate MPXV neutralizing titer data of participants that received at least one dose of MVA-BN intradermally. Titers for V4 are stratified first by prior smallpox vaccination then by HIV status. Colored/black bars on each group indicate the geometric mean titer. Serum neutralizing titers were measured by a fluorescence-based microneutralization assay using authentic clade IIb MPXV. Horizontal, black dashed line in each panel indicates the positivity threshold for neutralization (Fig. 2). B) Same as A, but for V5. Statistical testing conducted by Kruskal-Wallis test with Dunn’s method for multiple comparisons.

**Figure S4**

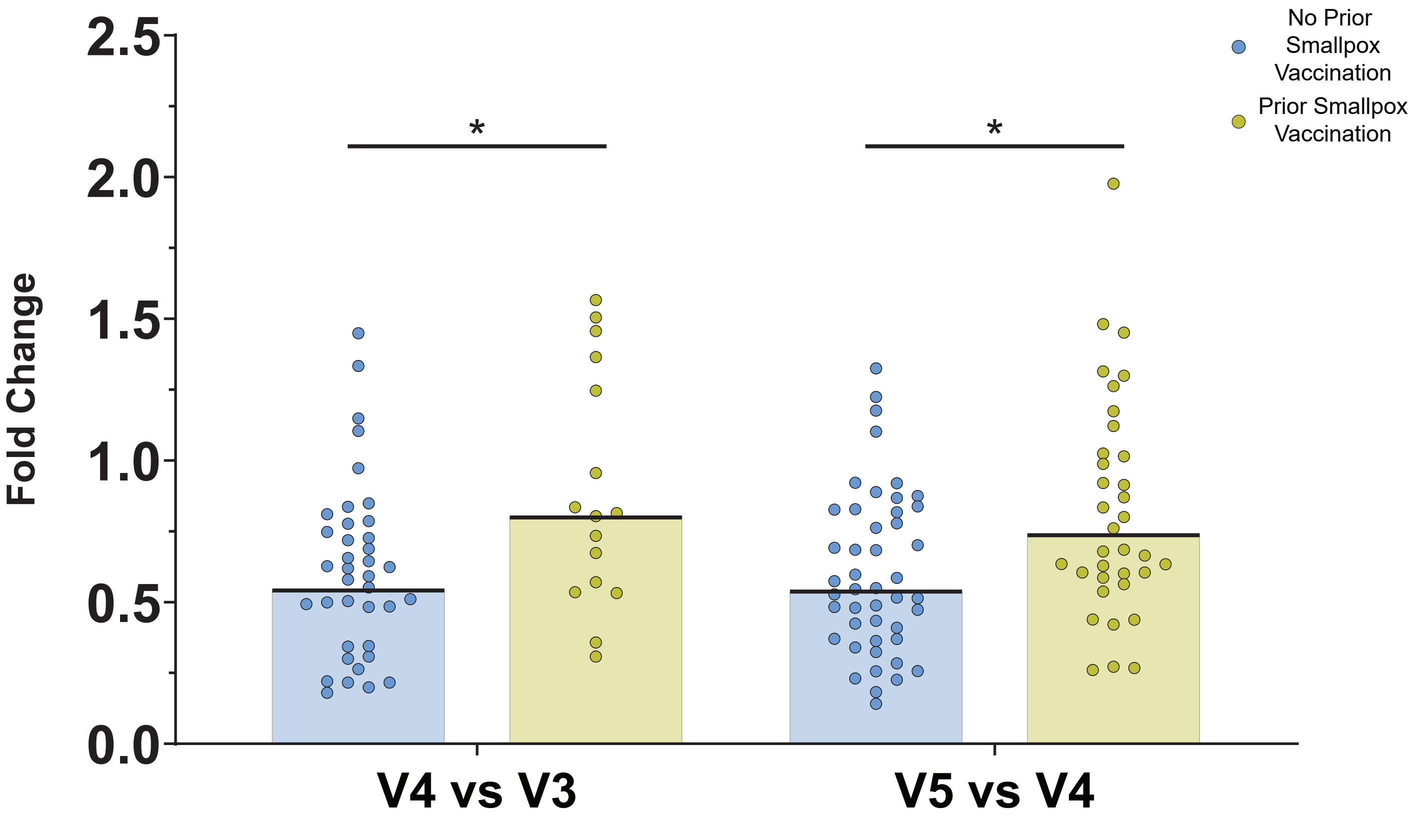
No Prior Smallpox Vaccination
Prior Smallpox Vaccination
2.5
2.0
1.5
1.0
0.5
0.0
Fold Change
*
*
V4 vs V3
V5 vs V4

**Figure S4: MPXV neutralizing titers wane significantly faster in naïve MVA-BN vaccinees.** Comparison of the paired fold changes in neutralizing titers from Fig. 2B and C. Colored/black bar for each group indicates the geometric mean fold change. Statistical testing conducted by Kruskal-Wallis test with Dunn's method for multiple comparisons.

### Figure S5

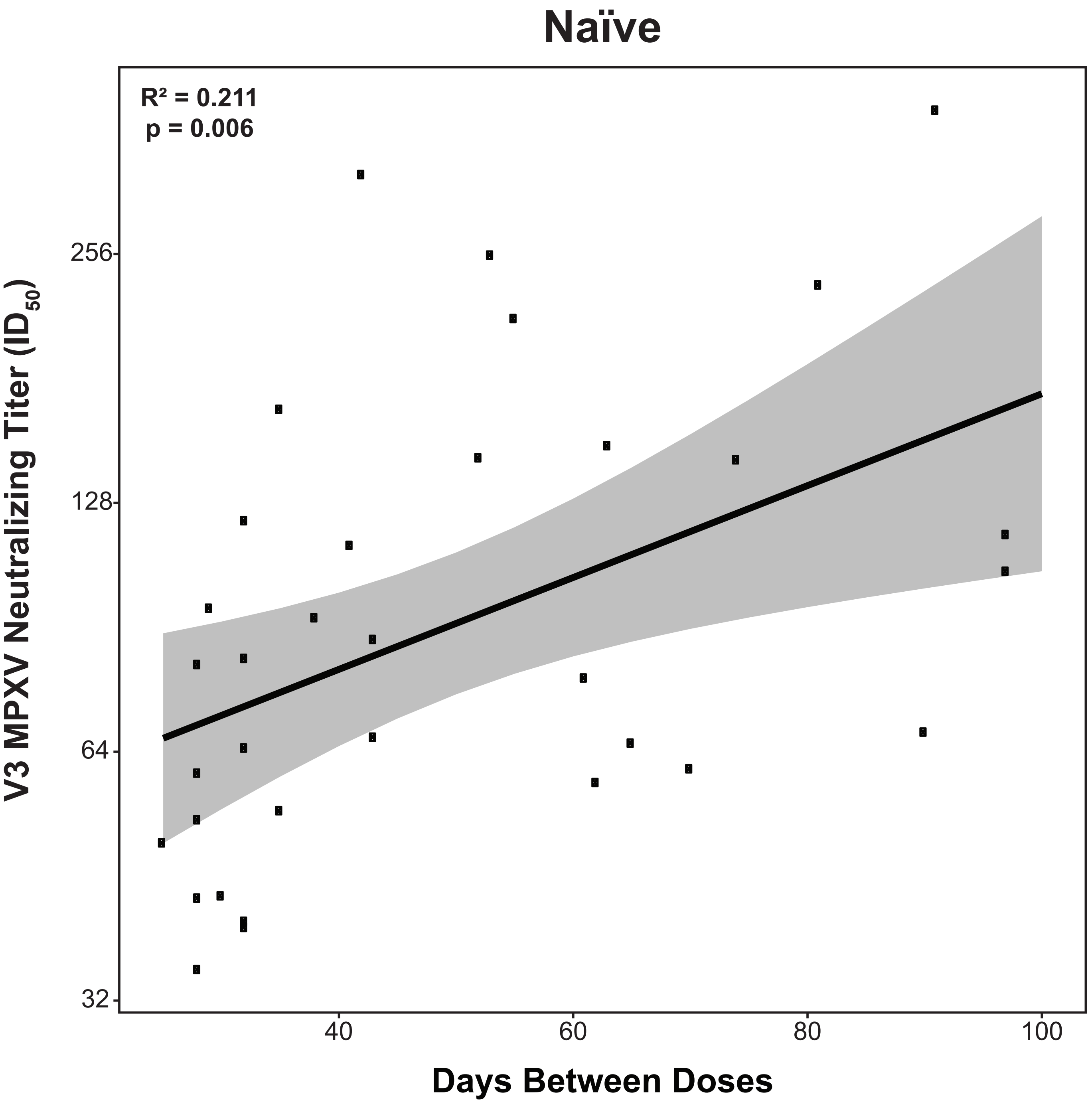

Naïve
R² = 0.211
p = 0.006
V3 MPXV Neutralizing Titer (ID50)
256
128
64
32
40
60
80
100
Days Between Doses

**Figure S5: Longer dosing intervals significantly correlate with peak MPXV neutralizing titers in naïve MVA-BN vaccinees.** Linear regression of V3 MPXV neutralizing titers versus dosing interval in VACV-naïve participants.

**Figure S6**

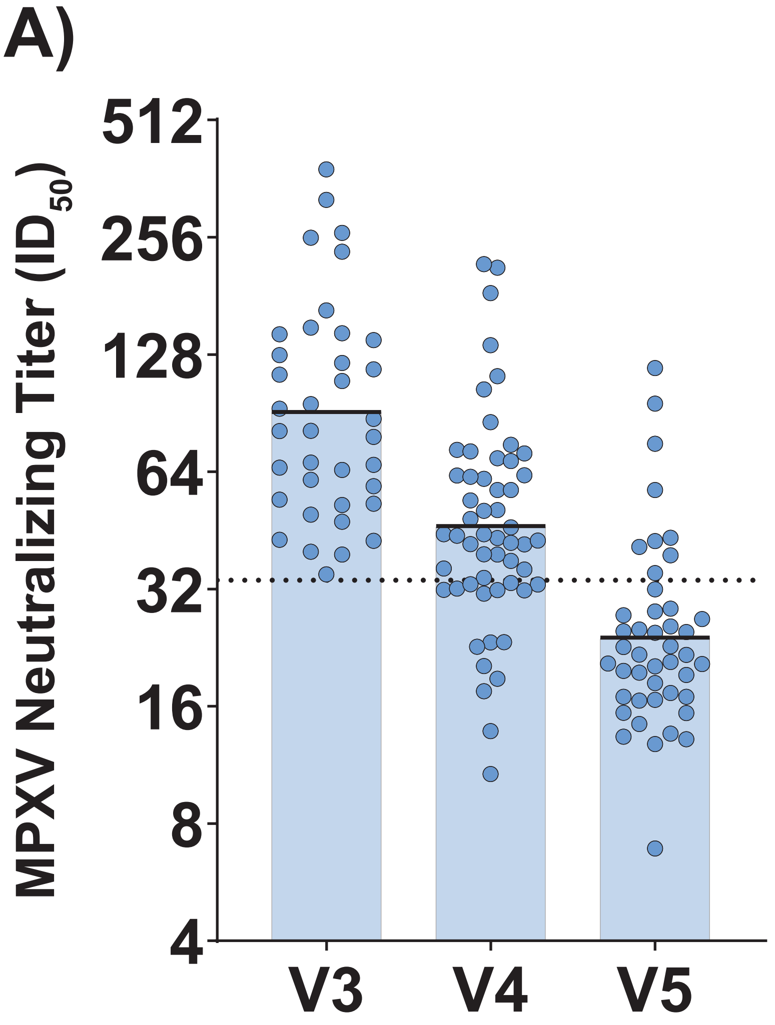


| | Naïve V3 | Naïve V4 | Naïve V5 |
|---|---|---|---|
| N | 37 | 54 | 44 |
| GMT (95% CI) | 91.1 (73.9-112.3) | 46.4 (39.2-54.9) | 24.0 (20.4-28.3) |
| Median DPV2 (IQR) | 19 (15-25) | 82 (72-97) | 294 (275-304) |
| % Neg | 0 | 29.6 | 81.8 |

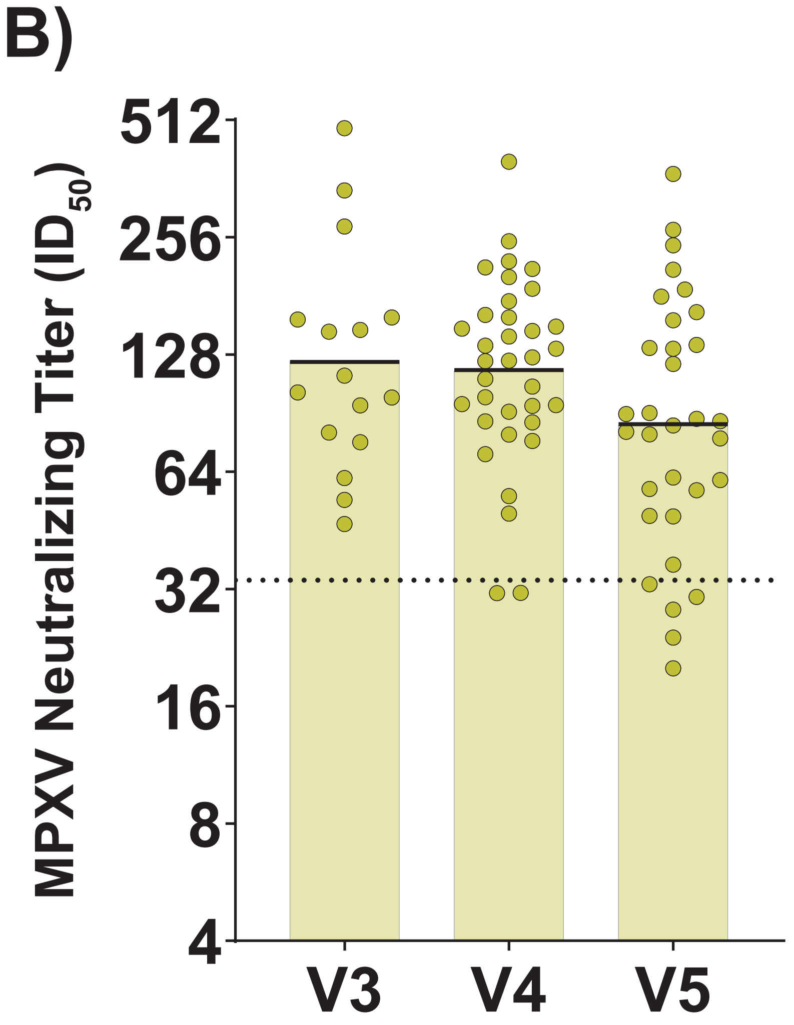


| | Exp V3 | Exp V4 | Exp V5 |
|---|---|---|---|
| N | 16 | 35 | 32 |
| GMT (95% CI) | 122.4 (86.5-173.3) | 116.7 (96.7-140.8) | 84.8 (65.0-110.7) |
| Median DPV2 (IQR) | 21 (18-29) | 81 (74-92) | 286 (267-313) |
| % Neg | 0 | 5.7 | 15.6 |

● No Prior Smallpox Vaccination

● Prior Smallpox Vaccination

**Figure S6: Subsets for multiplexed immunoassay analysis have similar neutralization to overall study groups.** A) Aggregate MPXV neutralizing titer data for naïve group subset used in Figures 4 and 5. Serum neutralizing titers were measured by a fluorescence-based microneutralization assay using authentic clade IIb MPXV. Colored/black bars on each group indicate the geometric mean titer. Horizontal, black dashed line indicates the positivity threshold for neutralization (Fig. 2). B) Same as A, but for experienced participants. Exp, experienced vaccinees (prior history of smallpox vaccination).

A)

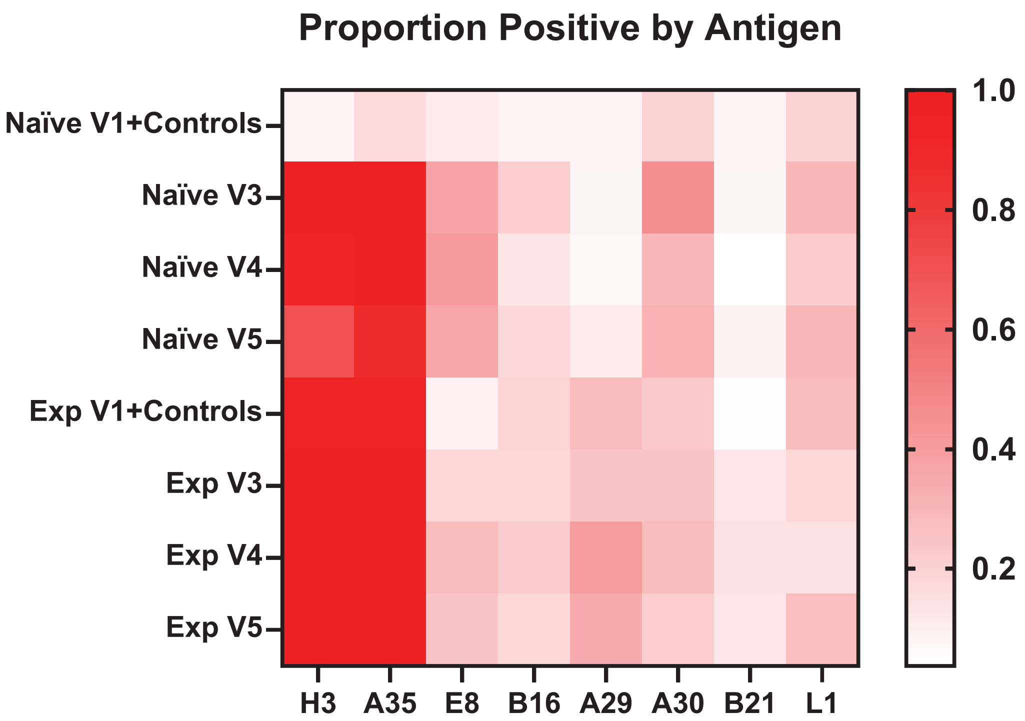
Proportion Positive by Antigen
Naïve V1+Controls
Naïve V3
Naïve V4
Naïve V5
Exp V1+Controls
Exp V3
Exp V4
Exp V5
H3
A35
E8
B16
A29
A30
B21
L1
1.0
0.8
0.6
0.4
0.2

B)

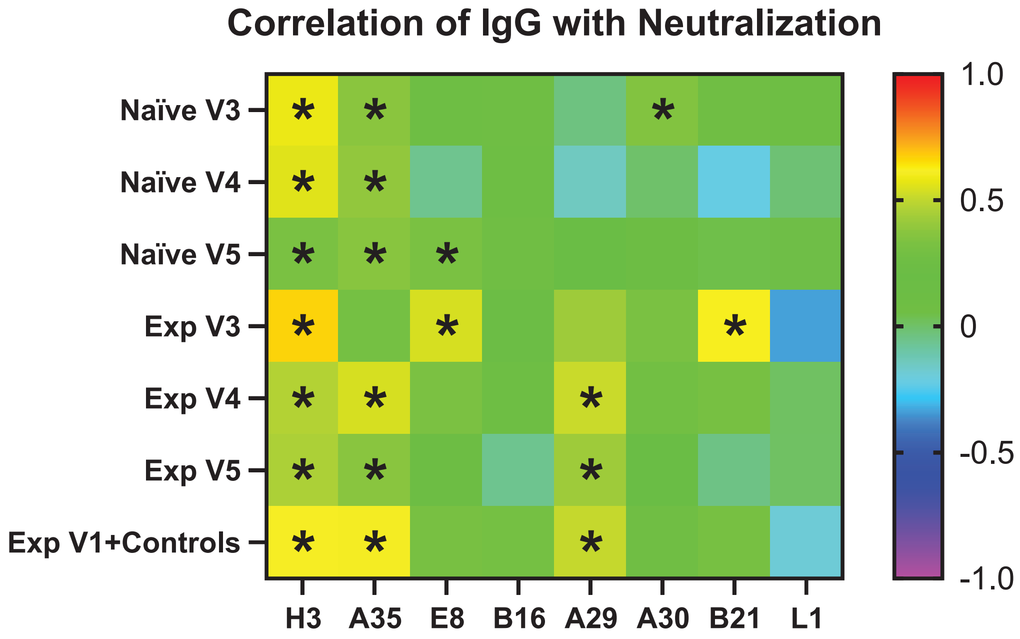
Correlation of IgG with Neutralization
Naïve V3
Naïve V4
Naïve V5
Exp V3
Exp V4
Exp V5
Exp V1+Controls
H3
A35
E8
B16
A29
A30
B21
L1
1.0
0.5
0
-0.5
-1.0

**Figure S7: MPXV H3 and A35 are immunodominant in all participants and significantly correlate with MPXV neutralization.** A) Proportion of participants that are IgG-positive for the indicated MPXV protein at a given time point and prior smallpox vaccination status. B) Spearman correlation for each MPXV protein against neutralizing titers by time point and prior smallpox vaccination status. Significant correlations, i.e., $p < 0.05$, are denoted with an asterisk. Exp, experienced vaccinees (prior history of smallpox vaccination).
